# Contribution of dominant and recessive model effects to the genetic architecture of Idiopathic Pulmonary Fibrosis

**DOI:** 10.64898/2026.02.18.26345897

**Authors:** Tamara Hernandez-Beeftink, Lauren J Donoghue, Abril Izquierdo, Samuel T Moss, Daniel Chin, Beatriz Guillen-Guio, Konain Fatima Bhatti, Simon Biddie, Nick Shrine, Richard Packer, Ayodeji Adegunsoye, Helen L Booth, CleanUP-IPF Investigators of the Pulmonary Trials Cooperative, William A Fahy, Tasha E Fingerlin, Ian P Hall, Simon P Hart, Mike R Hill, Nik Hirani, Naftali Kaminski, Elena Lopez-Jimenez, Jose Miguel Lorenzo-Salazar, Shwu-Fan Ma, Robin J McAnulty, Mark I McCarthy, Amy D Stockwell, Toby M Maher, Ann B Millar, Philip L Molyneaux, Maria Molina-Molina, Vidya Navaratnam, Margaret Neighbors, Justin M Oldham, Helen Parfrey, Gauri Saini, Ian Sayers, X Rebecca Sheng, Mary E Strek, Iain Stewart, Martin D Tobin, Moira KB Whyte, Yingze Zhang, Fernando J Martinez, Brian L Yaspan, Carl J Reynolds, David A Schwartz, Carlos Flores, Imre Noth, Alison E John, R Gisli Jenkins, Richard J Allen, Olivia C Leavy, Louise V Wain

## Abstract

**Rationale:** Idiopathic pulmonary fibrosis (IPF) is a rare, chronic, progressive lung disease with high mortality and few treatment options. Using an additive genetic model, genome-wide association studies (GWAS) have identified multiple risk loci highlighting new genes and pathways of interest. Since IPF risk could also be influenced by non-additive effects, we hypothesised that association analyses using alternative genetic models may provide additional mechanistic insight.

**Objectives:** To perform GWAS of IPF susceptibility to detect associations where the underlying effects are consistent with recessive or dominant genetic models.

**Methods:** We performed GWAS of IPF susceptibility, with logistic regression assuming dominant or recessive genetic models, including 5,159 IPF cases, from clinically-curated sources, and 27,459 controls. We functionally annotated independent signals and performed variant-to-gene mapping, applying fine-mapping to define potentially causal variants and genes. We assessed differential expression levels of genes of interest in publicly available single cell RNAseq data and in primary cells derived from IPF donors and controls.

**Main Results:** We identified five genome-wide significant signals, under a recessive model, that had not been reported previously. These included exonic variants in the cell-cycle gene Polyamine-Modulated Factor 1 (*PMF1*) and in Epsin 3 (*EPN3*) genes. We also observed evidence of increased *PMF1* expression in airway basal cells of IPF patients compared to controls.

**Conclusions:** Using alternative genetic models in IPF susceptibility GWAS identified new signals and genes, providing new insights into IPF pathogenesis and potential future therapies.

## INTRODUCTION

Idiopathic pulmonary fibrosis (IPF) is a rare chronic progressive illness of the respiratory system characterised by scarring of the lung interstitium (1)(2). Patients have a poor prognosis and currently there is a lack of treatments and therapeutic options that can stop or reverse the scarring of the lungs (1)(2)(3). The mean age at diagnosis is around 70 years of age, it is infrequent in individuals under 50 years old, and it is estimated to affect more than 7 per 100,000 people (4).

Multiple studies have focused on the identification of genetic risk factors associated with IPF (5)(6)(7)(8)(9)(10). The heritability of IPF due to single nucleotide polymorphisms (SNPs) has been estimated around 32% (11). Genome-wide association studies (GWAS) have identified multiple common signals of association and highlighted new genes and pathways of interest (5)(6)(7)(8)(9)(10), implicating genes involved in lung defence, telomere maintenance, cell-cell adhesion, cell proliferation, and fibrotic signalling.

All previously published IPF GWAS used an additive genetic model, which assumes an increase in risk for each copy of the effect allele. However, the genetic variation of complex traits could also be influenced by non-additive effects. GWAS using other genetic models such as the dominant model, where one or more copies of the effect allele are sufficient to increase risk, and the recessive model, where two copies are required to alter risk (12)(13), could provide new insight and a better understanding of the genetic variation in complex diseases (14), such as IPF.

Here, we hypothesised that association analyses assuming non-additive genetic models may identify new IPF susceptibility signals where the underlying effect is consistent with a recessive or dominant allelic effect.

## METHODS

### Study design and samples

We analysed seven independent case–control studies: Colorado (15), UUS (8), UK (7), IPF Job Exposures Study (IPF-JES) (16), Genentech (17), United States, United Kingdom, and Spain (US) (6) and Study of Clinical Efficacy of Antimicrobial Therapy Strategy Using Pragmatic Design in Idiopathic Pulmonary Fibrosis - University of California, Davis (CleanUP-UCD) (18) (**Figure 1**).

**Figure 1.**
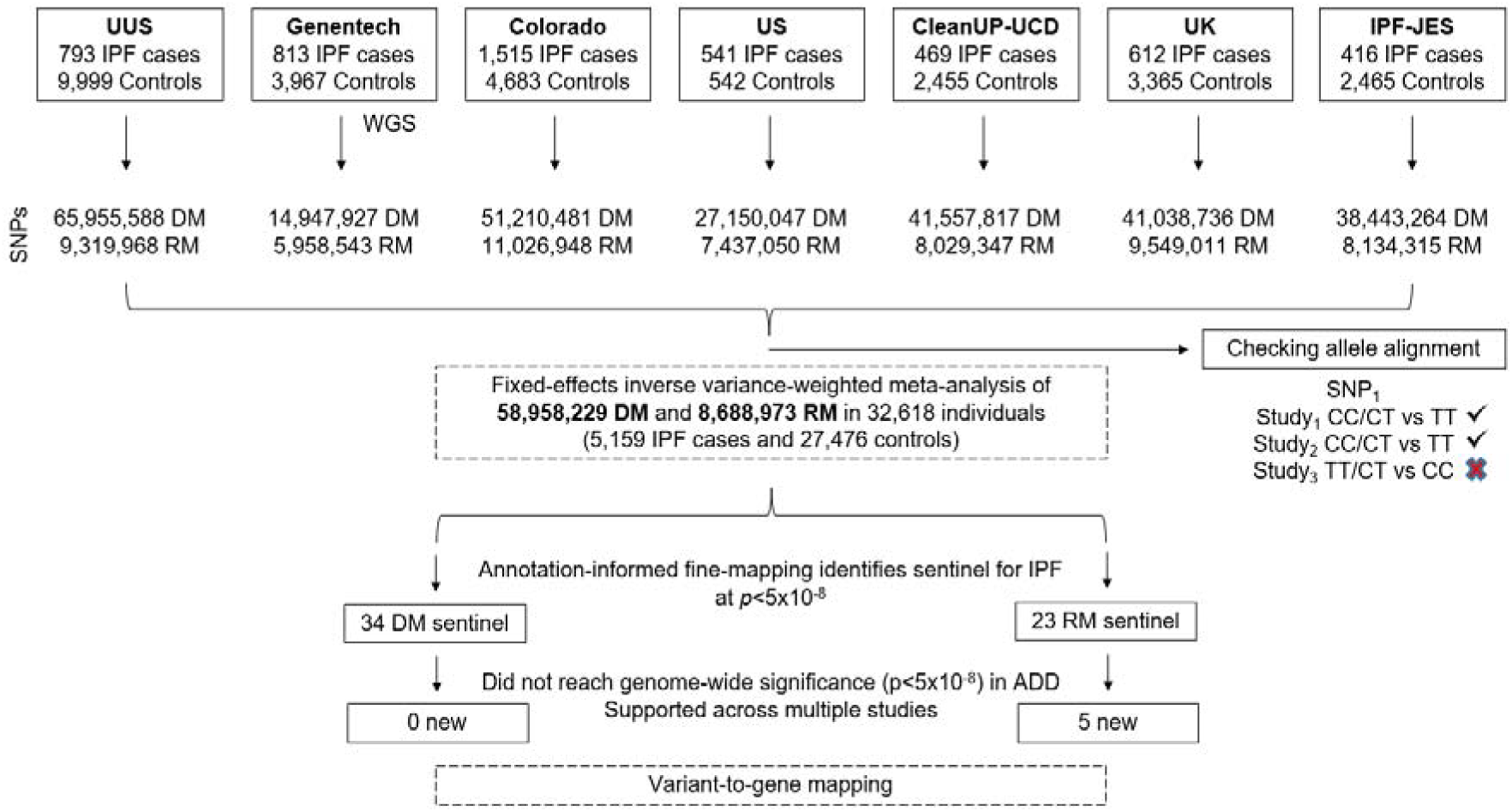
Sample selection workflow. SNPs: Single Nucleotide Polymorphisms; DM: Dominant genetic model; RM: Recessive genetic model; ADD: Additive genetic model. WGS: Whole-genome sequencing data.

All participating studies included unrelated individuals of genetically-determined European ancestry, with cases diagnosed according to the American Thoracic Society and European Respiratory Society guidelines (19). More information for each study can be found in the supplementary material. Written informed consent and ethics approval were correctly undertaken in all the studies following The Code of Ethics of the World Medical Association (Declaration of Helsinki) and approved by the appropriate institutional review or Research Ethics Committee.

### Genotyping and pre-processing

Genotyping arrays (Affymetrix and Illumina) were used to genotype individuals from the Colorado, IPF-JES, UK, US, UUS, and CleanUP-UCD studies. Sample quality control included the removal of individuals with low call rate estimates, sex-mismatched data, high heterozygosity rates, non-European genetic ancestry, had high relatedness, and duplicates across studies. These six studies were imputed using the TOPMed whole-genome sequencing (WGS) reference panel (GRCh38) via the TOPMed Imputation Server (20)(21)(10). For the Genentech study, genotypes were obtained from WGS using the HiSeq X Ten platform (Illumina) to an average read depth of 30X. Related individuals and those with a call rate of less than 10% were removed from the analysis. More details can be found in previous publication (10) and are summarised in the supplementary material.

### Association and statistical analysis

For six studies (Colorado, IPF-JES, UK, US, UUS, and CleanUP-UCD), genome-wide association analyses were conducted separately under dominant and recessive genetic models, using logistic regression with PLINK 2 (22) and adjusting each study for the first ten principal components to correct for population stratification. Variants for which the effect alleles were not aligned across all contributing studies were excluded from the meta-analysis. Variants with poor imputation quality (r^2^<0.5) were removed. In the Genentech study, dominant and recessive associations were performed using logistic regression with PLINK 1.9 (22), including sex, age, and five genetic-ancestry principal components as covariates.

The seven datasets were meta-analysed using inverse-variance weighted fixed effect meta-analysis in METAL (23), excluding variants that did not pass quality control in at least two studies. We estimated the genomic inflation factor using LDSC (24).

### Signal selection

Independent signals were defined based on a *p*-value threshold of *p*<5.0×10^-8^ (genome-wide significance) in ±500kb window. Independence from previously reported signals was determined based on genomic distance (±500 kb) and linkage disequilibrium (r²<0.1). We excluded signals where the association in any individual study was more significant than in the meta-analysis. The forest plot R v4.1.3 package was used to visualise the per-study and meta-analysis p-values. Significant signals were compared with the corresponding results, under an additive genetic model, using the same dataset (10). Whilst our primary genome-wide significance threshold was p<5×10^-8^, we also highlighted signals that met more conservative thresholds: i) a Bonferroni-corrected genome-wide significance threshold for two models (*p*<5×10^-8^ / 2), and ii) a more stringent threshold of *p*<5×10^-9^, used in previous large GWAS (25). This is because each variant was tested under two different (though not independent) genetic models, and TOPMed imputation enables testing of a larger number of variants than in previous GWAS. We compared the recessive and dominant model association results for all SNPs previously reported to be associated with IPF susceptibility at genome-wide significance using the additive model (*p*<5×10^-8^).

### Identification of putative causal genes and variants

#### Variant functional annotation

We annotated sentinel variants using Ensembl Variant Effect Predictor (VEP) v.115, and we identified potentially causal variants using the Wakefield Bayes factor method to calculate the posterior inclusion probability (PIP) of each variant to define a 95% credible set (26). To assess the regulatory potential and rank the functional roles of each SNP and its associated gene, we queried RegulomeDB (27), SpliceAI (28), and PrimateAI-3D (29). To map genetic variants within the 95% credible sets to alternative transcripts, we first annotated them to MANE-defined reference exons (Matched Annotation from NCBI and EMBL-EBI (30), as well as to Ensembl-annotated (Ensembl_EIC) and unannotated alternative exons (FLIbase_ENIC) (31). We then used FLIBase full-length transcript expression data derived from long-read mRNA sequencing (32) to assess expression of reference and alternative transcripts across tissue types. We plotted the transcripts for each related gene from FLIBase using the ‘ggtranscript’ function in R version 4.3.1.

We also annotated regions for functionality based on active chromatin marks, including DNase I hypersensitivity hotspots (DHS), transcription factor footprints, enhancer RNAs (eRNAs), Assay for Transposase-Accessible Chromatin using sequencing (ATACseq), ChIP sequencing (CHIPseq), and epigenetic modification of the DNA packaging protein histones (H3K27ac) (33). We used Open Targets to determine whether the variants had previously been associated with other phenotypes (*p*<0.001) (see supplementary material for further details). We conducted single-variant PheWAS analyses using a recessive model for 1,929 phenotypes in up to 393,063 European individuals for 389 quantitative traits and up to 158,054 cases and 394,247 controls for 1,540 binary traits in UK Biobank (UKBB) using the R Deep-PheWAS package (34).

#### Variant to gene mapping

Nearest-gene assignment was based exclusively on genomic distance, making it independent of prioritisation approaches that incorporate functional annotations. We followed the locus-to-gene (L2G) framework implemented in the Open Targets platform (https://platform-docs.opentargets.org/gentropy/locus-to-gene-l2g) (35), using the footprint distance metric to link variants to genes. According to this definition, a gene was considered the nearest one if the variant was located within its genomic footprint or if the linear distance to the transcriptional boundaries of the gene (TSS or transcription end site) was minimal. Gene coordinates were obtained from protein-coding canonical transcripts in Ensembl release 115. When a variant overlapped the coding regions of multiple canonical transcripts, all corresponding genes were reported and considered the nearest gene.

### Quantitative trait loci analysis

We analysed gene expression and splicing quantitative trait loci (eQTLs and sQTLs) across 54 tissues from the Genotype-Tissue Expression Project (GTEx) V8 (36) (downloaded in July 2020 from https://www.gtexportal.org/; data is only available under the additive model). We also accessed plasma protein expression quantitative trait loci (pQTLs) statistics from UKBB for 46,836 individuals from the general population (data available only under additive model). We used coloc (37) R v4.1.3 package to investigate whether the same variant was driving both the genetic association with IPF risk and an association with gene, protein, or splice variant expression levels, and report shared signals with a colocalization probability >0.7.

### Rare variant associations

We looked for exonic rare variant associations (minor allele frequency [MAF]<1%) with ‘pulmonary fibrosis’ within ±500 kb of our IPF sentinel variants using both single-variant and gene-based collapsing tests from 281,104 UKBB exomes (UKBB field IDs: 1121 & 22135; and ICD-10 code J841) accessed via the AstraZeneca PheWAS portal (https://azphewas.com/). We used a threshold of *p*<5×10^-6^ for both single-variant and gene-based tests.

### Nearby Mendelian respiratory disease and mouse knockout ortholog genes

We looked for rare Mendelian disease genes from ORPHANET (https://www.orpha.net/) and human orthologs of mouse knockout genes from the International Mouse Phenotyping Consortium (https://www.mousephenotype.org/) that were within ±500 kb of each IPF-associated sentinel and were linked with respiratory-related terms (‘fibrosis’, ‘fibrotic’, ‘lung’, ‘pulmonary, ‘respiratory’, ‘immune’, ‘inflammation’, ‘inflammatory’).

#### Cell culture

In order to investigate the differences between the basal epithelial cells of the airways in IPF and non-IPF lungs, primary airway basal epithelial cells were isolated from fresh human lung tissues obtained from lung cancer resection (non-IPF; n=3) or from lung explant tissue (IPF; n=3). All tissue samples were collected by the Clinical Research Facility Biobank at the Royal Brompton and Harefield Hospitals under ethical approval (NRES reference 20/SC/0142). Fresh tissue samples were cut into 1-2 mm^2^ diameter cubes and plated at 20-30 per dish in petri dishes coated with 3 ug/ml collagen (PureCol® Bovine Collagen, Cell Systems). The biopsies were left to adhere to petri dishes for 10 mins before adding 4 ml of airway epithelial cell growth media (Merck) supplemented with Bovine Pituitary Extract (0.004 ml/ml), rhEpidermal Growth Factor (10 ng/ml), rhInsulin (5 µg/ml), Hydrocortisone (0.5 µg/ml), Epinephrine (0.5 µg/ml), Triiodo-L-thyronine (6.7 ng/ml), rhTransferrin (10 µg/ml) and Retinoic Acid (0.1 ng/ml). Tissues were maintained for 7-10 days until at least 50% of the biopsies had released basal cells before removal to reduce fibroblast contamination. Cells were grown to confluence in a humidified incubator set to 37°C with 5% CO2 and passaged using TrypLE (Life Technologies). All isolated basal cells were confirmed positive for KRT5 by immunofluorescence staining and mRNA analysis.

#### RNA extraction and mRNA gene expression

Basal cells at passage 2 were grown to confluence and RNA extraction was performed using a Maxwell® RSC SimplyRNA Cell Kit according to the manufacturers protocol using Maxwell® RSC Extraction System (Promega). 100 ng of basal cell RNA from each sample was reverse transcribed into complimentary DNA (cDNA) using SuperscriptIV (Thermo Fisher Scientific) according to the manufacturer’s recommendations. cDNA was subjected to quantitative RT-PCR analysis using Fast SYBR™ green (Thermo Fisher Scientific) and gene specific primers for PMF1 and B2M (housekeeping gene) using the QuantStudio 3. Primers sequence were as follows: PMF1-F:AGCTACCAGAGATTCACTGACT; PMF1-R: AGATTTGCTGTGTCATCGCAG; B2M-F: AATCCAAATGCGGCATCT; B2M-R: GAGTATGCCTGCCGTGTG.

#### Integration of publicly available scRNAseq Data

Three single-cell RNA sequencing (scRNA-seq) datasets were integrated to create a large representative IPF scRNA sequencing dataset (see supplementary material for further details). The integrated datasets were: GSE136831 (38), GSE135893 (39), and GSE128033 (40), obtained from NCBI’s public data repository. The integrated scRNA dataset included 52 IPF and 48 control samples. Non-IPF ILD samples were removed from two of the datasets (GSE136831 and GSE135893) during data processing and quality control. Cell-level filtering included removing cells with mitochondrial ratios <20% and retaining cells with >500 unique molecular identifiers per cell and >1,000 genes per cell. Differential expression of genes across different cell types at Human Lung Cell Atlas (HLCA) level 3 annotation was generated using R packages ggplot2 and Seurat. Statistical significance was calculated using the ‘t.test’ function in R (ggsignif package extension).

## RESULTS

We analysed data from 5,159 IPF cases and 27,459 controls of European ancestry, and a total of 59 million and 8.7 million SNPs with high imputation quality for the dominant and recessive genetic models, respectively (**Figure 2; Table S1; Figure S1**). In the recessive model, the lower number of SNPs reflects the minimum number of minor allele homozygote genotypes required for analysis. We identified 57 independent significant signals (34 using the dominant model and 23 using the recessive model). Of these, five had not been previously reported, and were supported across multiple studies (**Table 1**; **Figure 3; Table S2-S3; Figure S2-S3**). These five signals were detected only when using a recessive genetic model. Of those, four met a more stringent threshold accounting for testing of two models (p<2.5×10^-8^), and two of those four also met a more stringent genome-wide threshold of p<5×10^-9^.

**Figure 2.**
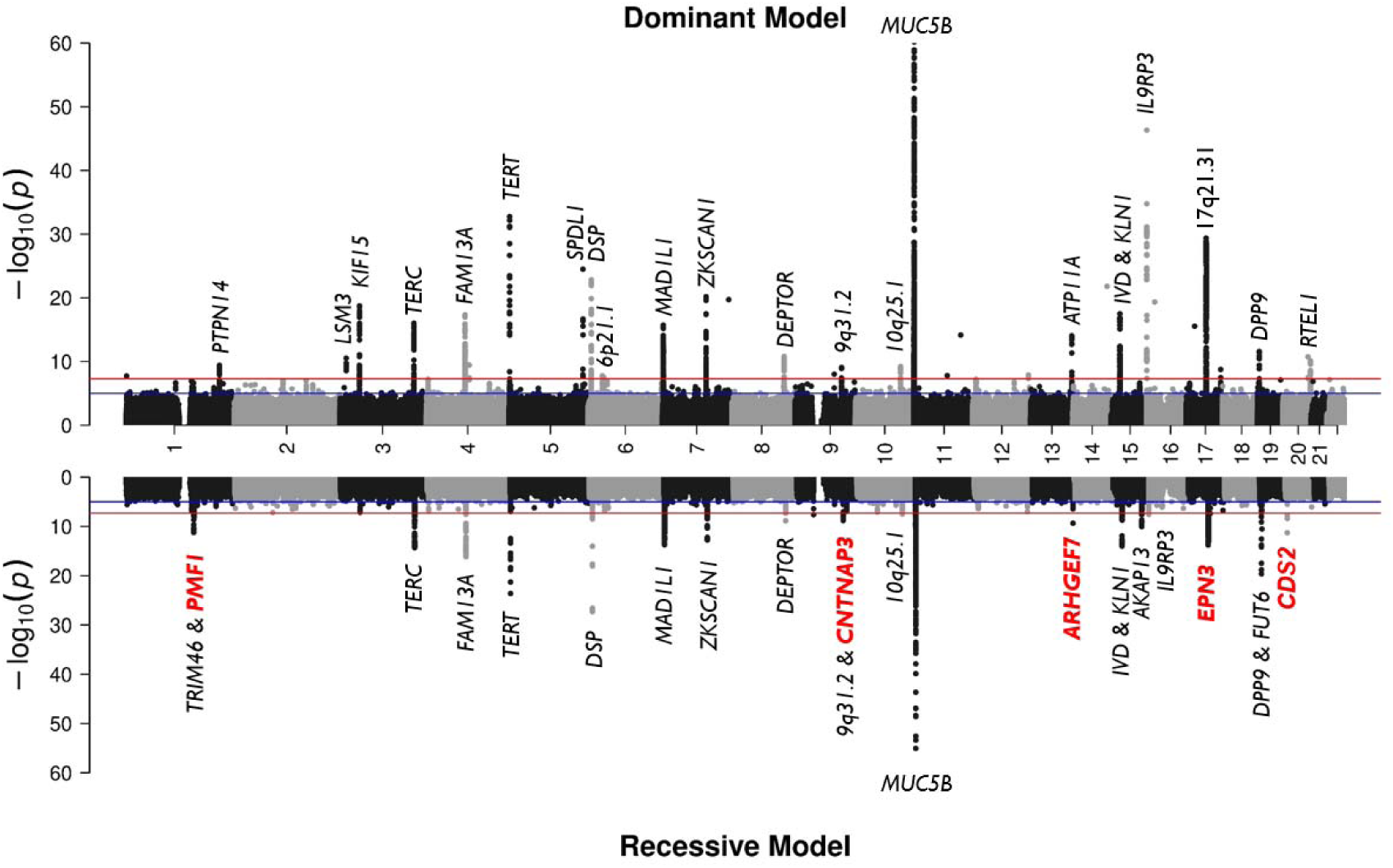
Mirror Manhattan plot of meta-analysis results from the dominant (up) and recessive (down) genetic model. The y-axis shows the transformed *p*-values (–log10) while the x-axis represents chromosomal coordinates (GRCh38). The horizontal red line corresponds to the genome-wide threshold (*p*=5×10^-8^) and the blue line shows the suggestive significance threshold (*p*=5×10^-5^). The genomic inflation factor of the meta-analysis results from the dominant (λ=1.05) and recessive (λ=1.06) genetic models did not show major deviations from the null hypothesis of no association. The plot was truncated at *p*=10^-60^. Newly discovered annotated nearest genes are highlighted in red and bold. The most significant previously reported *MUC5B* signal (rs35705950) has a p<5×10^-121^ in both genetic models.

**Figure 3.**
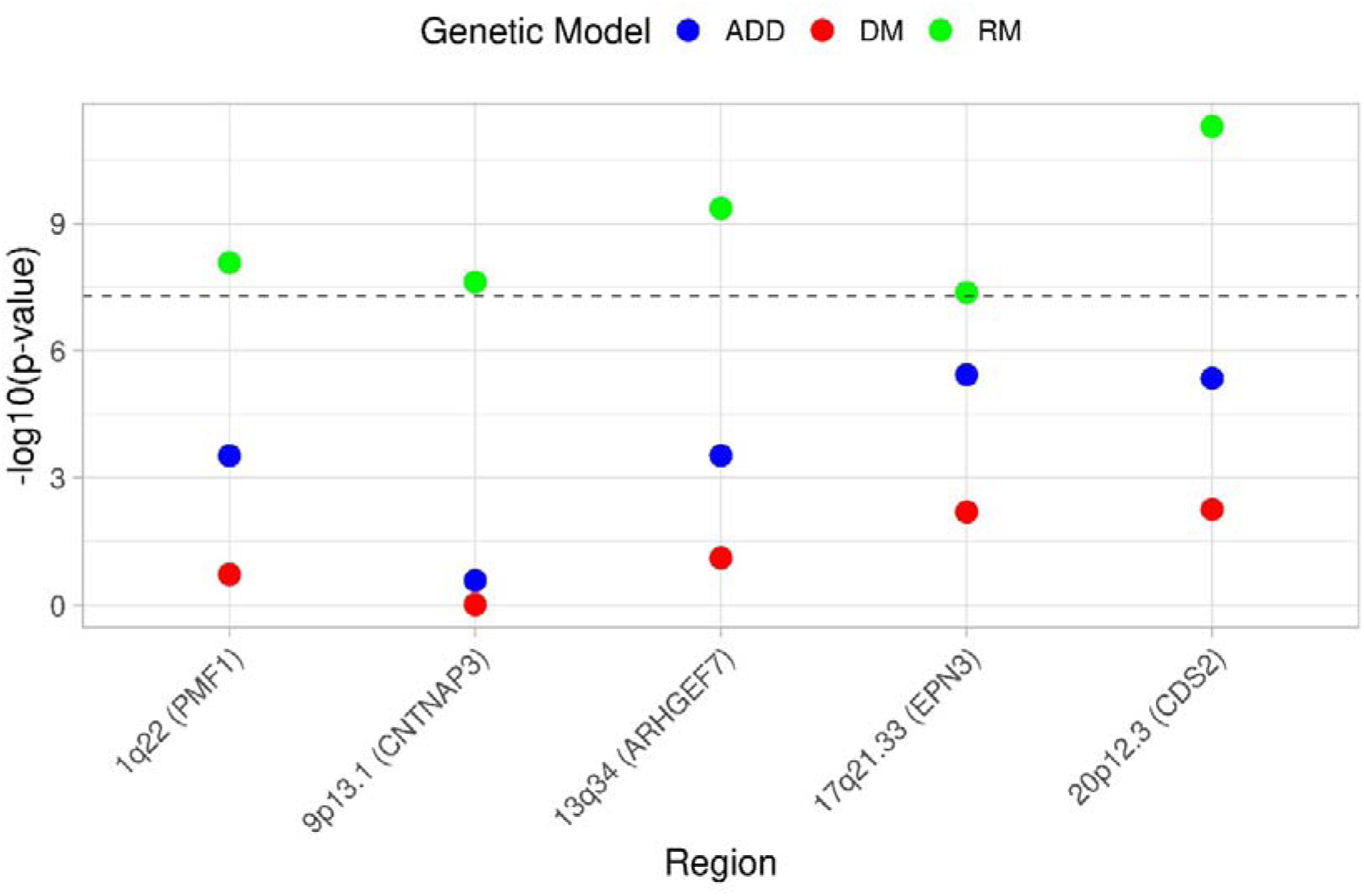
Summary plot of the 5 new significant signals in the non-additive genetic models. The x-axis shows the region with nearby genes harbouring the obtained SNPs. The y-axis shows the transformed *p*-values (-log10). The horizontal dashed line corresponds to the genome-wide significance threshold (*p*=5×10^-8^). Each colour represents the transformed *p*-value obtained from the different genetic models: in blue the additive results (ADD), in red the results of the dominant genetic model (DM) and in green the recessive genetic model results (RM).

**Table 1.** Five new signals of association with IPF detected when assuming a dominant or recessive genetic model.

|  |  |  |  |  |  | Additive Genetic Model |  |  | Dominant Genetic Model |  | Recessive Genetic Model |  |  |  |
| --- | --- | --- | --- | --- | --- | --- | --- | --- | --- | --- | --- | --- | --- | --- |
| CHR | BP | SNP | Location | Nearest gene | A1/A2 | A1 Freq. | OR (CI95%) | P | OR (CI95%) | P | OR (CI95%) | P | Significant model effect allele | No. SNPs in credible set (largest PIP) |
| 1 | 156232382 | rs1052053 | exonic | *PMF1* | G/A | 0.371 | 0.91 (0.87-0.95) | $3.00 \times 10^{-4}$ | 0.96 (0.90-1.02) | 0.189 | 0.75 (0.68-0.83) | $8.38 \times 10^{-9}$ | (GG vs AA;AG) | 22 (0.164) |
| 9 | 38912426 | rs74455724 | intergenic | *CNTNAP3* | A/G | 0.190 | 1.05 (0.96-1.15) | 0.259 | 1.00 (0.91-1.10) | 0.962 | 2.08 (1.61-2.70) | $2.40 \times 10^{-8}$ | (AA vs AG;GG) | 2 (0.857) |
| 13 | 111161343 | rs11838956 | intronic | *ARHGEF7* | T/C | 0.241 | 1.11 (1.05-1.17) | $2.97 \times 10^{-4}$ | 1.06 (1.00-1.13) | 0.077 | 1.47 (1.30-1.66) | $4.40 \times 10^{-10}$ | (TT vs TC;CC) | 1 (0.997) |
| 17 | 50541888 | rs4794159 | exonic | *EPN3* | A/C | 0.449 | 1.12 (1.07-1.18) | $3.67 \times 10^{-6}$ | 1.11 (1.04-1.19) | $6.34 \times 10^{-3}$ | 1.24 (1.15-1.33) | $4.27 \times 10^{-8}$ | (AA vs AC;CC) | 2 (0.905) |
| 20 | 5245010 | rs6116709 | intergenic | *CDS2* | C/T | 0.389 | 1.18 (1.10-1.27) | $4.46 \times 10^{-6}$ | 1.17 (1.05-1.29) | $6.00 \times 10^{-3}$ | 1.62 (1.41-1.86) | $5.17 \times 10^{-12}$ | (CC vs CT;TT) | 1 (0.995) |

A1: effect allele; A2: non-effect allele; Freq: Frequency; OR: Odds Ratio; PIP: posterior inclusion probability; Coordinates in GRCh38; CI: Confidence Interval.

Bayesian fine-mapping was performed for each of the five loci to identify the most likely causal variants (**Table S3**). For the chromosome 1 (sentinel rs1052053), 22 variants formed the 95% credible set, two of which had a PIP>0.1 (rs1052053 in an exon of Polyamine-Modulated Factor 1 (*PMF1),* PIP=0.164, and rs35002119 in an intron of the same gene, PIP=0.150). For the other four signals, the 95% credible sets each contained a single SNP with PIP>0.85 (two credible sets contained only a single SNP with PIP=1.00). These were: an intergenic SNP between *CNTNAP3* and *FAM201A*, an intergenic SNP between *CDS2* and *PROKR2*, an intronic SNP in *ARHGEF7*, and an exonic SNP in *EPN3*. All sentinel variants were common (MAF>5%). None of the signals had nearby mouse ortholog knockout genes related to respiratory traits, nor did they overlap with any gene-based or rare-variant associations in the UKBB or with protein levels in blood.

*PMF1* encodes polyamine modulated factor 1, which is involved in chromosome segregation (41). The sentinel exon variant (rs1052053-G) is a missense variant resulting in a glutamine to arginine substitution at position 75 (Gln75Arg), with a combined annotation-dependent depletion (CADD) score of 15.72 (implying it is amongst the 10% most harmful variants in the genome). Three variants, including the sentinel variant, in the 95% credible set (rs1052053, rs2241108, and rs2853641) overlap exons of *PMF1* in the reference transcript (MANE; ENST00000359511), which is expressed in all tissues (**Figure S4**). This signal (chromosome 1 sentinel rs1052053) is also associated with the expression and splicing of multiple other genes across multiple tissues (**Table S3**). In addition, one variant with a very low contribution to the credible set (rs2072499, PIP=0.006) may affect splicing of *SLC25A44* and is present in an exon of four alternative transcripts of *SLC25A44*: two expressed in all tissues, one expressed at low levels in 13 tissues, and one specific to pancreas and prostate (**Figure S4**).

The sentinel rs1052053-G had been previously associated at genome-wide significance with reduced cystatin C, serum urea, and creatinine levels (42) and increased risk of stroke (43). The PheWAS analysis using a recessive model highlighted strong associations primarily with renal biomarkers (**Figure S5**). Of note, rs1052053-G has a lower frequency (∼30%) in individuals of African ancestry compared with other ancestries (35-40%; The 1000 Genomes Project (1KGP)). Furthermore, a rare third allele (C) has been detected at a frequency of 0.08% in European populations (source: The 1KGP) but this was not analysed in this study. Analysis by ORPHANET revealed that the *LAMTOR2*, *LMNA* and *RIT1* genes, located within ±500 kb of rs1052053 were also implicated in rare Mendelian immune or respiratory diseases (**Table S3**). Analysis of publicly available scRNA-seq data revealed significantly increased *PMF1* gene expression in fibroblasts and basal cells from IPF patients compared with controls (**Figure S6**), with decreased expression in Alveolar Type II (AT2) cells. We observed a potential increased *PMF1* expression in basal cells from IPF patients using qPCR in primary cells isolated from fresh human lung tissue (**Figure S7**).

The exonic signal (rs4794159) in *EPN3,* encoding Epsin 3 which has a role in endocytosis, was annotated as ‘deleterious’. The risk allele (rs4794159-A) leads to a proline to threonine amino acid substitution at position 544 (Pro544Thr). This signal was also associated with the expression and splicing of several other genes across multiple tissues (**Table S3**). The second variant in the 95% credible set, rs7215760 (PIP=0.088), also is located in an exon in one alternative transcript that was expressed across different tissue types, including low-level expression in lung (**Figure S4)**. At the single-cell level, *EPN3* was expressed in basal cells of individuals with IPF but was not detected in controls (**Figure S6**). This signal (rs4794159) had not been associated with other traits at genome-wide significance, although it was nominally associated with sex hormone binding protein levels (*p*=5×10^-6^) (44). Rare variants in the nearby gene *ITGA3* have previously been implicated in a multi-organ disorder interstitial lung disease-nephrotic syndrome-epidermolysis bullosa (ILNEB) syndrome (**Table S3**).

The third signal was an intronic variant to *ARHGEF7* (Rho Guanine Nucleotide Exchange Factor 7). *ARHGEF7* expression was increased in basal cells and fibroblasts from IPF patients compared with controls (**Figure S6**). The two new intergenic association signals were located nearest the *CNTNAP3* gene on chromosome 9 and the *CDS2* gene on chromosome 20. The signal on chromosome 9 was associated with expression of *IGFBPL1* and *TCEA1P3* in the brain (**Table S3**).

We also evaluated the effects of 35 signals previously reported for additive association with IPF susceptibility. All were most significant under an additive model, with the exception of the signals at the *TRIM46, DSP,* and *FUT6* loci, which were more significant under a recessive model (*p_additive_*= 1.53×10^-9^ vs *p_recessive_*= 9.78×10^-12^; *p_additive_*=4.81×10^-53^ vs *p_recessive_*= 9.43×10^-64^; *p_additive_*= 1.05×10^-9^ vs *p_recessive_*= 3.06×10^-11^; respectively) (**Table S4; Figure S8**).

## DISCUSSION

We identified five new genetic signals of association with IPF susceptibility that would not have been identified when using an additive model in the same dataset and adds to accumulating evidence for altered mitotic spindle assembly in IPF, new evidence for endocytosis processes, and new evidence for cellular membrane trafficking. This highlights the value of considering alternative genetic models in GWAS for IPF.

The new exonic signal on chromosome 1 implicates the *PMF1* gene both through the presence of a missense variant (Glutamine > Arginine) within the signal, and an association with *PMF1* gene expression in lung tissue with increased expression observed in basal cells of IPF patients compared with controls. Interestingly, variants in linkage disequilibrium with rs1052053 have been implicated in alternative splicing of *PMF1* in lung tissue and in COVID-19 severity (45). However, rs1052067, which is associated with COVID-19 severity (r^2^=0.630 with rs1052053), was not within our fine-mapped 95% credible set. *PMF1* encodes Polyamine Modulated Factor 1, which forms part of the MIS12 complex, one of the multi-protein complexes that constitute the kinetochore that connects chromosomes to spindle microtubules to enable chromosome segregation during mitosis (46). Recent GWAS of IPF susceptibility have also implicated additional mitotic spindle assembly complex genes, including *SPDL1* (47), *KNL1* (9), *KIF15* and *MAD1L1* (46). *KIF15,* which encodes Kinesin Family Member 15 and promotes spindle elongation by sliding microtubules apart, has been shown to be down-regulated in replicating epithelial cells from IPF patients (48). This suggests that epithelial cells in the IPF lung experience impaired regenerative capacity, with reductions in both proliferation and appropriate differentiation (49), alongside an increase in the proportion of cells entering a senescent state (50)(51). Of note, a recent plasma genome-wide Mendelian randomization study identified methylation levels of *PMF1* as a potential causal influence on IPF risk (52).

We also observed a new exonic signal on chromosome 17 implicating the *EPN3* gene. This gene encodes an endocytic adaptor protein involved in clathrin-mediated endocytosis and cellular membrane trafficking (53). Studies have shown that endocytosis is disrupted in fibrosis (54), and inhibition of endocytosis alters collagen I homeostasis (55). Clathrin-mediated endocytic pathways can also regulate TGF-β receptor signalling (56). When this signalling is dysregulated, it promotes persistent fibroblast activation and differentiation into myofibroblasts, as well as excessive extracellular matrix deposition. Neither *PMF1* nor *EPN3* are currently under investigation as a therapeutic targets at any stage of clinical drug development.

Another new signal on chromosome 13 was an intronic variant to *ARHGEF7*, a guanine nucleotide exchange factor for Rho family GTPases. This is functionally similar to *AKAP13* (A-kinase anchoring protein 13), another guanine nucleotide exchange factor previously associated with IPF (7). The nearest genes to the other new association signals were *CNTNAP3* (Contactin Associated Protein Family Member 3), which encodes a cell recognition protein, and *CDS2* (Cytidine Diphosphate Diacylglycerol Synthase 2), a key enzyme in phosphoinositide biosynthesis involved in cell signaling, migration, and angiogenesis. We were unable to confirm variant-to-gene mapping for these non-coding signals despite utilising multiple *in silico* approaches.

The power to detect non-additive effects in GWAS of complex traits is lower than that for additive effects (57). We maximised statistical power by including all seven available, clinically-curated IPF case-control datasets in our discovery stage. In the absence of a further dataset with adequate power for independent replication, we utilised a pragmatic approach to identify robust signals that included evaluation of per-study p-values compared to meta-analysis p-values to ensure that only signals that were supported across independent datasets, and not driven by individual studies, were reported. We also highlighted signals that met more conservative significance thresholds. However, independent replication across additional datasets remains the gold standard for assessing the robustness and generalisability of association signals. A lack of publicly available QTL data generated under recessive and dominant models limits our ability to further assess whether variant-gene expression, splicing, and protein level associations also align with a dominant or recessive model. To maintain consistency with our recessive-model GWAS, we conducted a recessive-model PheWAS, recognising that this model provides less statical power than an additive-model but may be more likely to detect a true recessive effect on gene expression. Our datasets comprised individuals of European genetic ancestry; therefore these association signals need to be assessed in other populations to understand their generalisability.

In conclusion, the use of alternative genetic models identified new genome-wide significant signals which implicate new genes in IPF development. These findings add to recent mechanistic insights from other GWAS highlighting the importance of the mitotic spindle assembly complex and impaired cell proliferation in the pathogenesis of IPF.

## ETHICS

This research was conducted with appropriate ethics approval. The PROFILE study (which provided samples for the UK and UUS studies) had institutional ethics approval at the University of Nottingham (NCT01134822 - ethics reference 10/H0402/2) and Royal Brompton and Harefield NHS Foundation Trust (NCT01110694 - ethics reference 10/H0720/12). UK samples were recruited across multiple sites with individual ethics approval (University of Edinburgh Research Ethics Committee [The Edinburgh Lung Fibrosis Molecular Endotyping (ELFMEN) Study NCT04016181] 17/ES/0075, NRES Committee South West - Southmead, Yorkshire and Humber Research Ethics Committee 08/H1304/54 and Nottingham Research Ethics Committee 09/H0403/59). Spanish samples were recruited under ethics approval by ethics committee from the Hospital Universitario N.S. de Candelaria (reference of the approval: PI-19/12). The UUS study also included individuals from clinical trials with ethics approval (ACE [NCT00957242] and PANTHER [NCT00650091]). For the UCSF cohort, sample and data collection were approved by the University of California San Francisco Committee on Human Research and all patients provided written informed consent. For the Vanderbilt cohort, the Institutional Review Boards from Vanderbilt University approved the study and all participants provided written informed consent before enrolment. For individuals recruited at the University of Chicago, consenting patients with IPF who were prospectively enrolled in the institutional review board-approved ILD registry (IRB#14163A) were included. Individuals recruited at the University of Pittsburgh Medical Centre had ethics approval from the University of Pittsburgh Human Research Protection Office (referenceSTUDY20030223: Genetic Polymorphisms in IPF). Individuals from the COMET (NCT01071707) and Lung Tissue Research Consortium (NCT02988388) studies were also included in the Chicago study. All subjects in the Colorado study gave written informed consent as part of IRB-approved protocols for their recruitment at each site and the GWAS study was approved by the National Jewish Health IRB and Colorado Combined Institutional Review Boards (COMIRB). Subjects in the Genentech study provided written informed consent for whole-genome sequencing of their DNA. Ethical approval was provided as per the original clinical trials (INSPIRE [NCT00075998], RIFF [NCT01872689], CAPACITY [NCT00287729 and NCT00287716] and ASCEND [NCT01366209]). Individuals in the CleanUP-UCD study included individuals from clinical trials with ethics approval (NCT02759120). These samples were genotyped under University of Virginia ethics approval (IRB 20845). IPFJES involved human participants and was approved by East Midlands - Nottingham 1 Research Ethics Committee REC reference: 17/EM/0021IRAS project ID: 203355. Participants gave informed consent to participate in the study before taking part.

## DATA AVAILABITILY

Summary statistics (i.e., effect size estimates, standard errors, *p* values and basic variant information) for all variants included in the genome-wide meta-analysis can be accessed via https://github.com/genomicsITER/PFgenetics.

## AUTHORS CONTRIBUTION

LVW, OCL and RJA designed and supervised the study. TH-B, LJD, AI, SM, DPC, BG-G, KFB, SB, NS, and RP performed the analyses. AA, HLB, CleanUP-IPF Investigators of the Pulmonary Trials Cooperative, WAF, TEF, IPH, SPH, MRH, NH, SJ, NK, EL-J, JML-S, S-FM, RJM, MIM, ADS, TMM, ABM, PLM, MM-M, VN, MN, JMO, HP, GS, IS, XRS, MES, MDT, MKBW, YZ, FM, BLY, CJR, DAS, CF, IN, AEJ, RGJ, and LVW participated in data collection. TH-B, and LVW wrote the first draft of the manuscript. All authors revised and approved the final version of the manuscript.

