## Supplementary material for "Contribution of dominant and recessive model effects to the genetic architecture of Idiopathic Pulmonary Fibrosis"

Indices

[**Table S2.** Five genome-wide signals from a dominant and recessive genetic models, detailed in each individual study [Excel spreadsheet].	6](#_Toc220495211)

[**Table S3.** Functional assessment of sentinels and variants included in the 95% credible set [Excel spreadsheet].	6](#_Toc220495212)

[**Table S4**. Sentinel previously reported genome-wide signals under an additive genetic model and results for a dominant and recessive genetic models [Excel spreadsheet].	6](#_Toc220495213)

**SUPPLEMENTAL METHODS**

#### **Study samples**

We conducted an analysis from seven independent case-control studies on idiopathic pulmonary fibrosis (IPF) previously documented. For all seven studies, IPF cases were diagnosed based on the guidelines established by the American Thoracic Society and European Respiratory Society. The seven datasets are composed by Colorado (1), Chicago (2), UK (3), IPF Job Exposures Study (IPF-JES) (4), Genentech (5) (6), United States, United Kingdom, and Spain (UUS) (7), and Study of Clinical Efficacy of Antimicrobial Therapy Strategy Using Pragmatic Design in Idiopathic Pulmonary Fibrosis - University of California, Davis (CleanUP-UCD) (8).

The Colorado study (1), a cohort of 1,515 patients with fibrotic idiopathic interstitial pneumonias (fIIP) was sourced from various US cohorts (including National Jewish Health IIP population, InterMune IPF trials, UCSF, Vanderbilt University IIP population, and the National Heart Lung and Blood Institute Lung Tissue Research Consortium). These cases were matched with 2,455 population controls chosen for their genetic similarity. Genotyping for all individuals was conducted using the Illumina Human 660W Quad BeadChip array.

The US study (previously referred as "Chicago" study) (2) encompassed 541 IPF cases from the University of Chicago, University of Pittsburgh, and the COMET study. It also included 542 population controls from the database of genotypes and phenotypes (dbGaP) and the University of Pittsburgh. Genotyping for both cases and controls was carried out using the Affymetrix Genome-Wide Human SNP 6.0 array.

The UK study (3) gathered a total of 612 IPF cases from 9 different centres across the UK, and 3,366 matching controls selected from the UK Biobank. Cases were genotyped with the Affymetrix UK BiLEVE array, while controls were genotyped with the UK Biobank array.

The IPF-JES (4) included 416 men from England, Scotland, or Wales diagnosed with IPF, along with 2,465 matching male controls selected from both the IPF-JES study and the UK Biobank. Genotyping was carried out using the Affymetrix UK Biobank array.

The Genentech study (5) (6) comprised 813 cases derived from three IPF clinical trials (ASCEND, CAPACITY, and RIFF) and 3,949 controls from four non-IPF clinical trials including age-related macular degeneration, diabetic macular edema, multiple sclerosis, asthma, and inflammatory bowel disease. Genotyping was accomplished through whole-genome sequencing using the HiSeq X Ten platform (Illumina) with an average read depth of 30X. Individuals were filtered based on ADMIXTURE v1.23 estimates to keep only the European individuals, and also by missingness, relatedness and heterozygosity. Variants were filtered according to missingness > 0.1, Hardy-Weinberg equilibrium (HWE) (p<=5x10^-8^), case-control missingness and allele balance.

The UUS study (United States, United Kingdom, and Spain) (7) included 793 IPF cases from 7 study cohorts (ACE, PANTHER, UCD, Chicago, UCSF, PROFILE, and Spain), along with 9,999 population controls selected from the UK Biobank to match ancestry, sex, and smoking distribution. Genotyping for cases was done using the Affymetrix UK Biobank and Spanish Biobank arrays, while controls were genotyped with the UK Biobank array.

The CleanUP-UCD study (8) encompassed a total of 469 IPF cases drawn from a randomized clinical trial conducted across 35 locations in the United States and from the University of California Davis. Additionally, 2,455 population controls were selected from the UK Biobank. Genotyping was performed on both cases and controls using the Affymetrix UK Biobank array.

Six case-control studies (Colorado, IPF-JES, UK, US, UUS, and CleanUP-UCD) were imputed using the TOPMed WGS reference panel (GRCh38) via the TOPMed Imputation Server, ensuring that only SNPs available in both (cases and controls) were included.

#### **Statistical analysis**

Bayesian fine-mapping

Fine mapping was used to generate a set of genetic variants with a 95% probability of containing the causal variant associated with a given genetic signal (95% credible set). This analysis was performed using R version 4.2.1. The fine-mapping strategy used, based on Wakefield approximate Bayes factor (9), assumed the existence of a single measured causal variant.

To assess the association between genetic variants within the 95% credible sets and gene expression and splicing in 49 tissues (including both lung and non-lung tissues), we used the GTEx portal.

eQTL, sQTL and pQTL colocalisation

For those variants showing associations with gene expression quantitative trait loci (eQTLs) and splicing quantitative trait loci (sQTLs), colocalisation analyses were performed in different tissues, including lung, cultured fibroblasts, and whole blood from (GTEx). These analyses were performed using the coloc v5.2.3 package (10) in R version 4.2.1.

Colocalisation analyses were used to determine whether the same causal variant was associated with both IPF susceptibility and gene expression or splicing in GTEx. The posterior probability for each model was estimated using the approximate Bayes factor for the following cases:

- H0: neither IPF susceptibility nor gene expression/splicing have a genetic association in that region
- H1: only IPF susceptibility has a genetic association in that region
- H2: only gene expression/splicing has a genetic association in that region
- H3: both IPF susceptibility and gene expression/splicing are associated, but with different causal variants
- H4: both IPF susceptibility and gene expression/splicing are associated and share a single causal variant

If the posterior probability in favour of the alternative hypothesis, indicating the presence of a common single causal variant for both IPF susceptibility and gene expression/splicing (H4), exceeded 70%, we inferred that the IPF and gene expression/splicing signals showed colocalisation.

For protein quantitative trait loci (pQTLs), we accessed summary statistics from the UK Biobank (UKBB) for 46,836 individuals from the general population with plasma protein expression quantitative trait loci (pQTLs) (additive model available). We also colocalised IPF signals with pQTLs using the same methods and procedures described above. For each signal, gene pairs were selected if there were significant pQTLs (p<5x10^-6^).

#### **Functional annotation**

We conducted a thorough functional impact assessment using empirical data from several integrated software tools and datasets. In addition, we used OpenTargets (<https://www.opentargets.org/>) (11) to access cross-referencing of genetic variants with a wide range of phenotypes. Variants were annotated for variant type, protein function, and pathogenic potential using Ensembl Variant Effect Predictor (VEP) (Ensembl release 115, GRCh38, including MANE Select and canonical transcripts).

#### **scRNAseq data integration**

Log-normalisation was performed on each dataset used for integration to adjust for technical noise and removal of batch effects due to differences in sequencing depth, followed by variance stabilisation transformation and identification of highly variable genes using negative binomial regression. Harmony (v0.1) was used to integrate multi-dataset cells into clusters (13).

The cell type annotation of the integrated dataset was conducted using Human Lung Cell Atlas (HLCA) (14) which consists of over two million cells from the respiratory tract of 486 individuals and combines together 49 different datasets to give a comprehensive platform for single-cell level annotation (15). There are five annotation levels in HLCA with level 1 (four cell types) being basic to level 5 being finest (53 cell types), that highlight different granularities of cell type identification. HLCA level 3 (25 cell types) was used in this study to show scRNA gene expressions.

### **SUPPLEMENTAL TABLES**

#### **Table S1.** Sample size of the study.

| Cohort | **Total** | **Cases** | **Controls** | **Autosomal variants analysed in the dominant model** | **Autosomal variants analysed in the recessive model** |
| --- | --- | --- | --- | --- | --- |
| Colorado | 6,198 | 1,515 | 4,683 | 51,210,481 | 11,026,948 |
| US | 1,083 | 541 | 542 | 27,150,047 | 7,437,050 |
| UK | 3,977 | 612 | 3,365 | 41,038,736 | 9,549,011 |
| IPF-JES | 2,881 | 416 | 2,465 | 38,443,264 | 8,134,315 |
| Genentech | 4,762 | 813 | 3,949 | 14,947,928 | 5,958,540 |
| UUS | 10,792 | 793 | 9,999 | 65,955,588 | 9,319,967 |
| CleanUP-UCD | 2,924 | 469 | 2,455 | 41,557,817 | 8,029,347 |
| **Total** | **32,618** | **5,159** | **27,459** | **58,958,229** | **8,688,973** |
| The number of individuals and variants shown is the result after quality controls. | | | | | |

#### **Table S2.** Five genome-wide signals from a dominant and recessive genetic models, detailed in each individual study [Excel spreadsheet].

#### **Table S3.** Functional assessment of sentinels and variants included in the 95% credible set [Excel spreadsheet].

#### **Table S4**. Sentinel previously reported genome-wide signals under an additive genetic model and results for a dominant and recessive genetic models [Excel spreadsheet].

### **SUPPLEMENTAL FIGURES**

**Figure S1.** Quantile-quantile plot of the observed versus expected -log10 p-values of the meta-analysis results from the dominant (left) and recessive (right) genetics models. The genomic inflation factor of the meta-analysis results from the dominant (λ=1.05) and recessive (λ=1.06) genetic models did not show major deviations from the null hypothesis of no association.

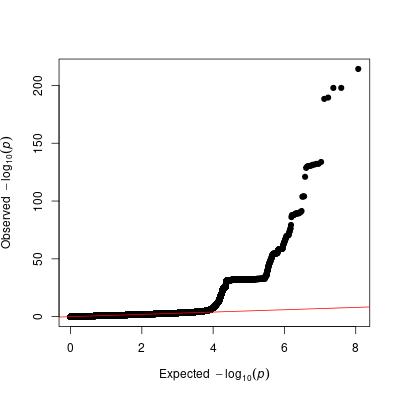

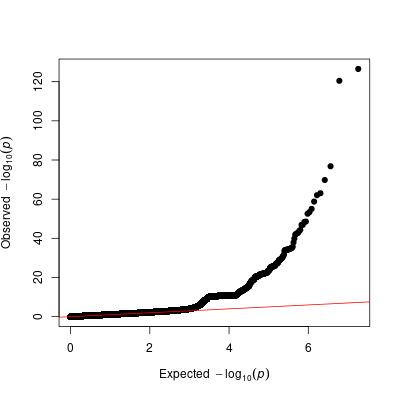

**Figure S2.** Forest plots of the signals of interest**:**

a) rs1052053 (*PMF1*)

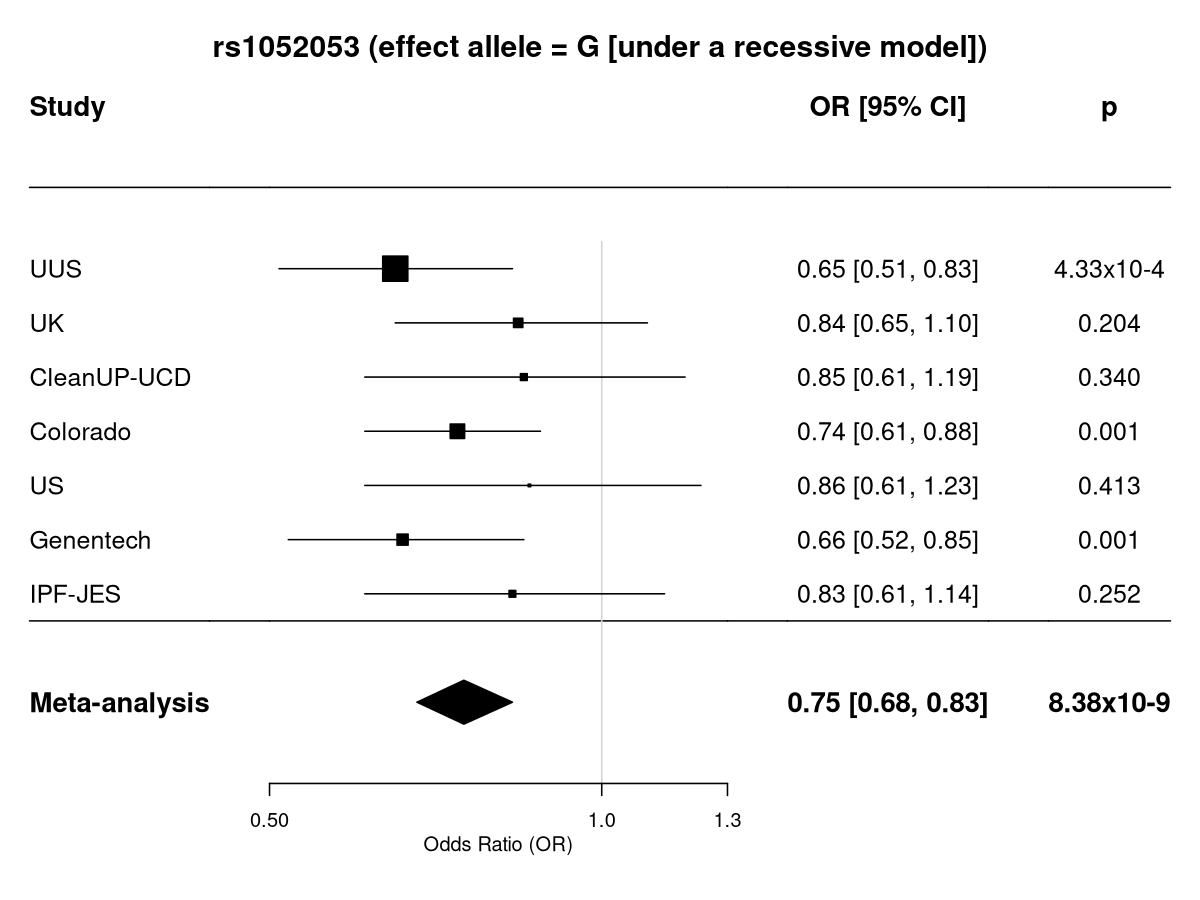

b) rs74455724 (*CNTNAP3*)

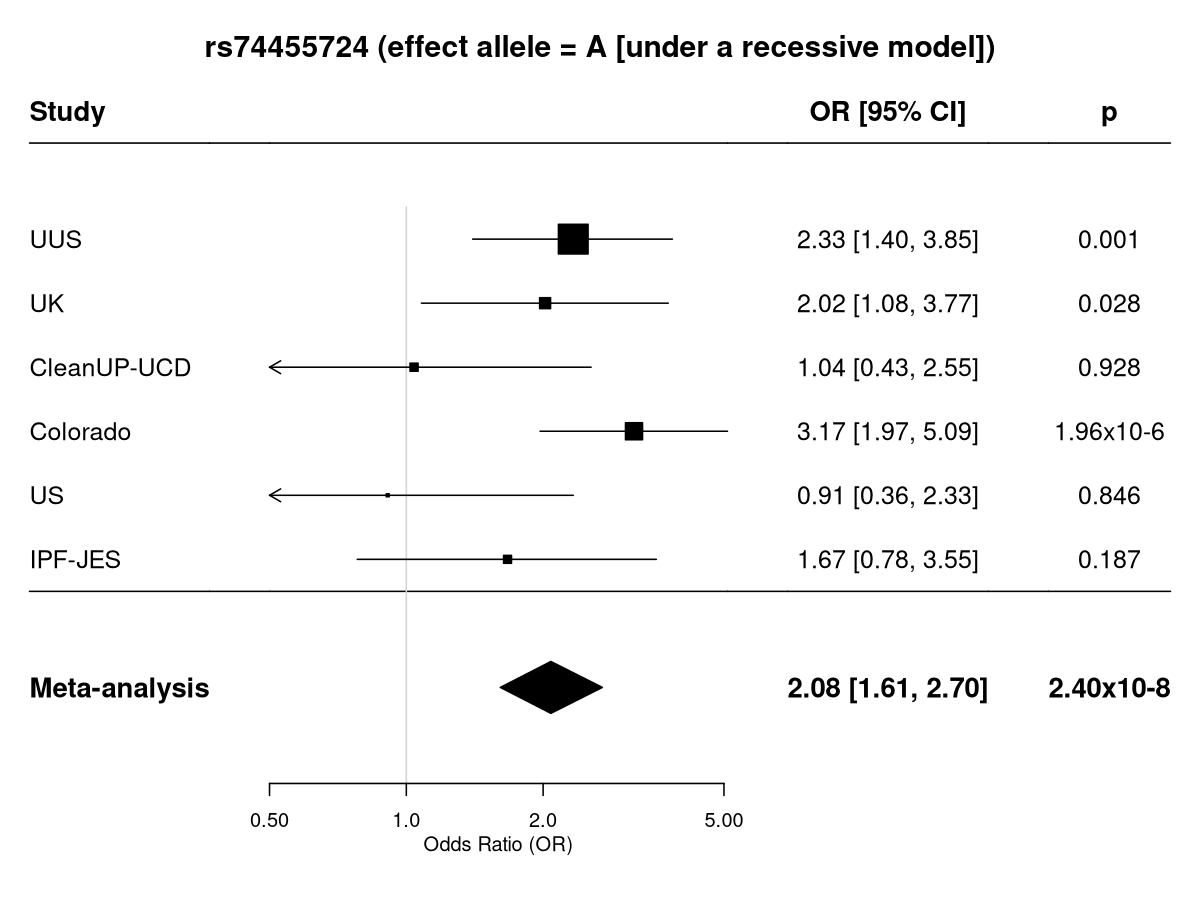

c) rs11838956 (*ARHGEF7*)

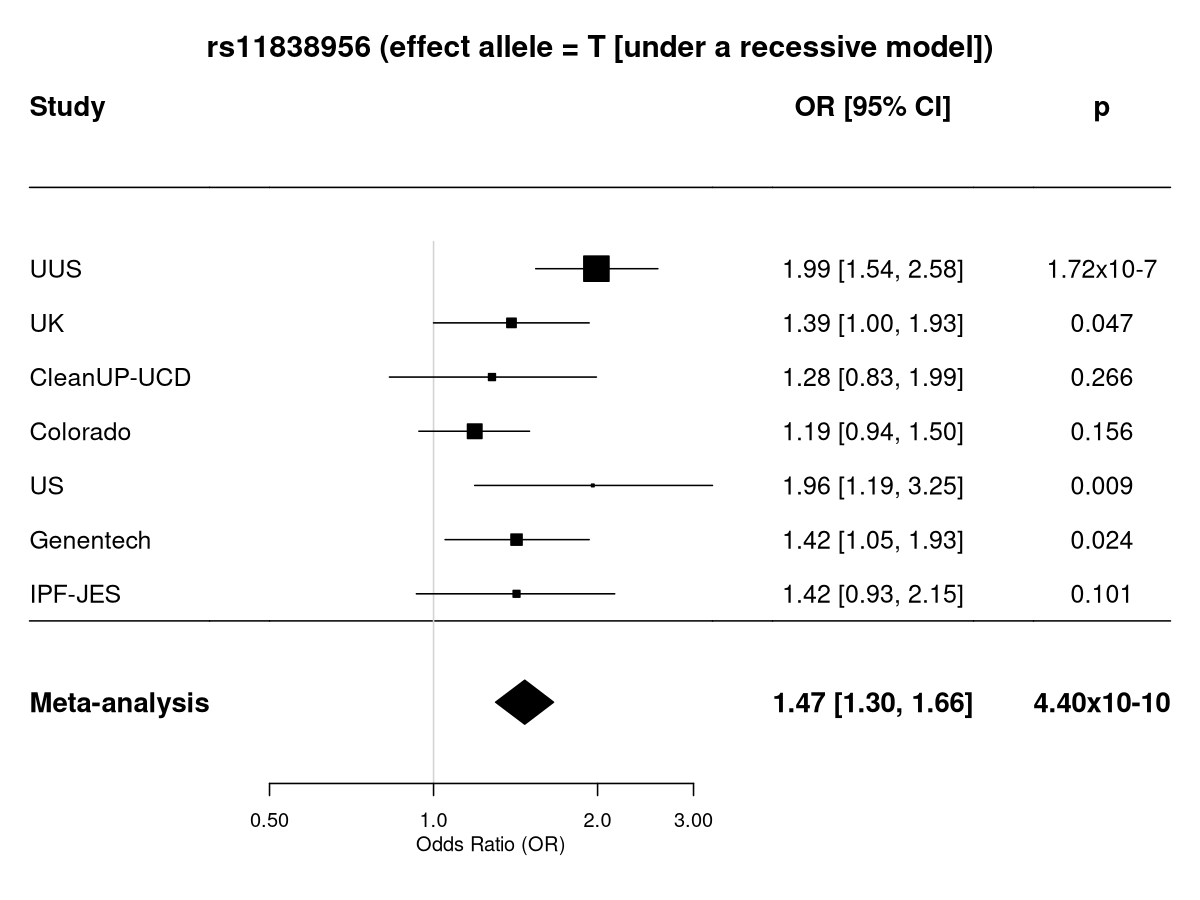

d) rs4794159 (*EPN3*)

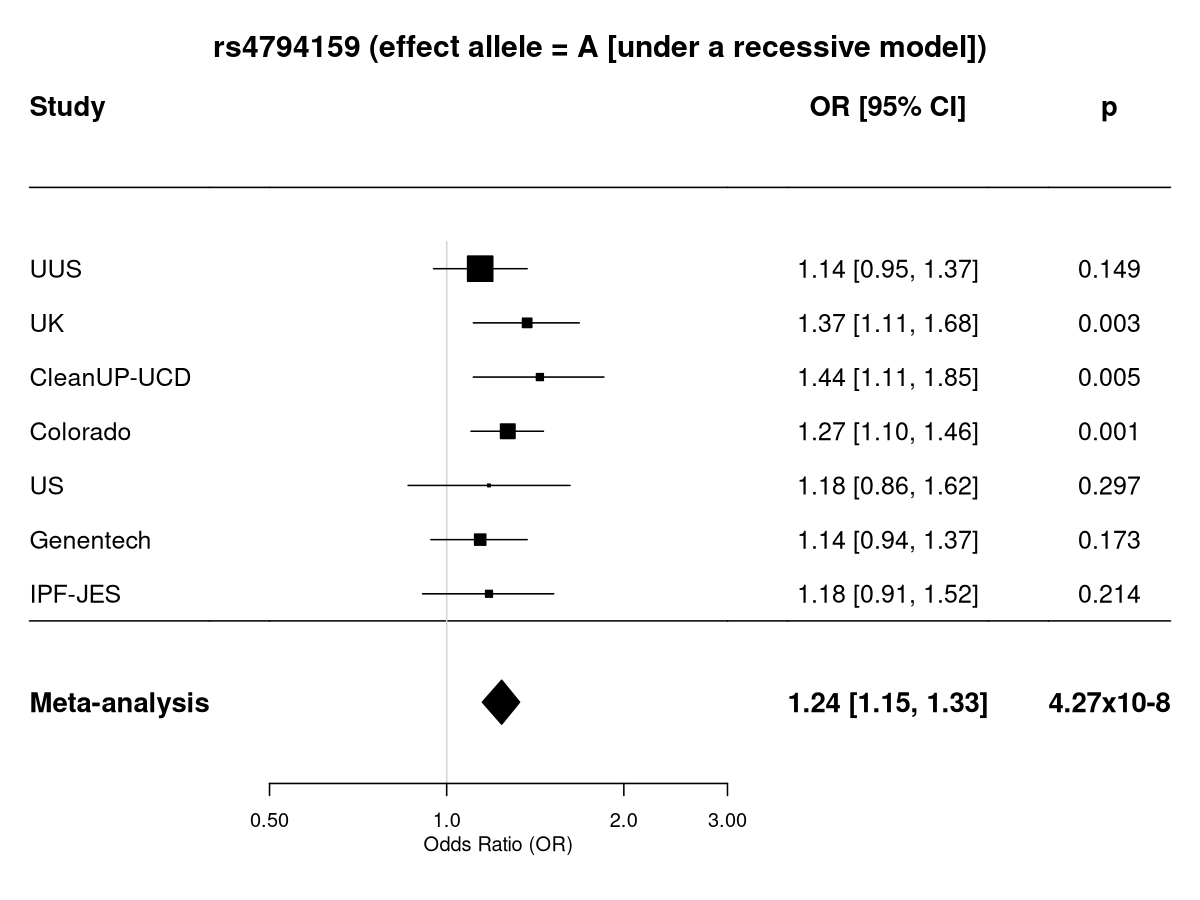

e) rs6116709 (*CDS2*)

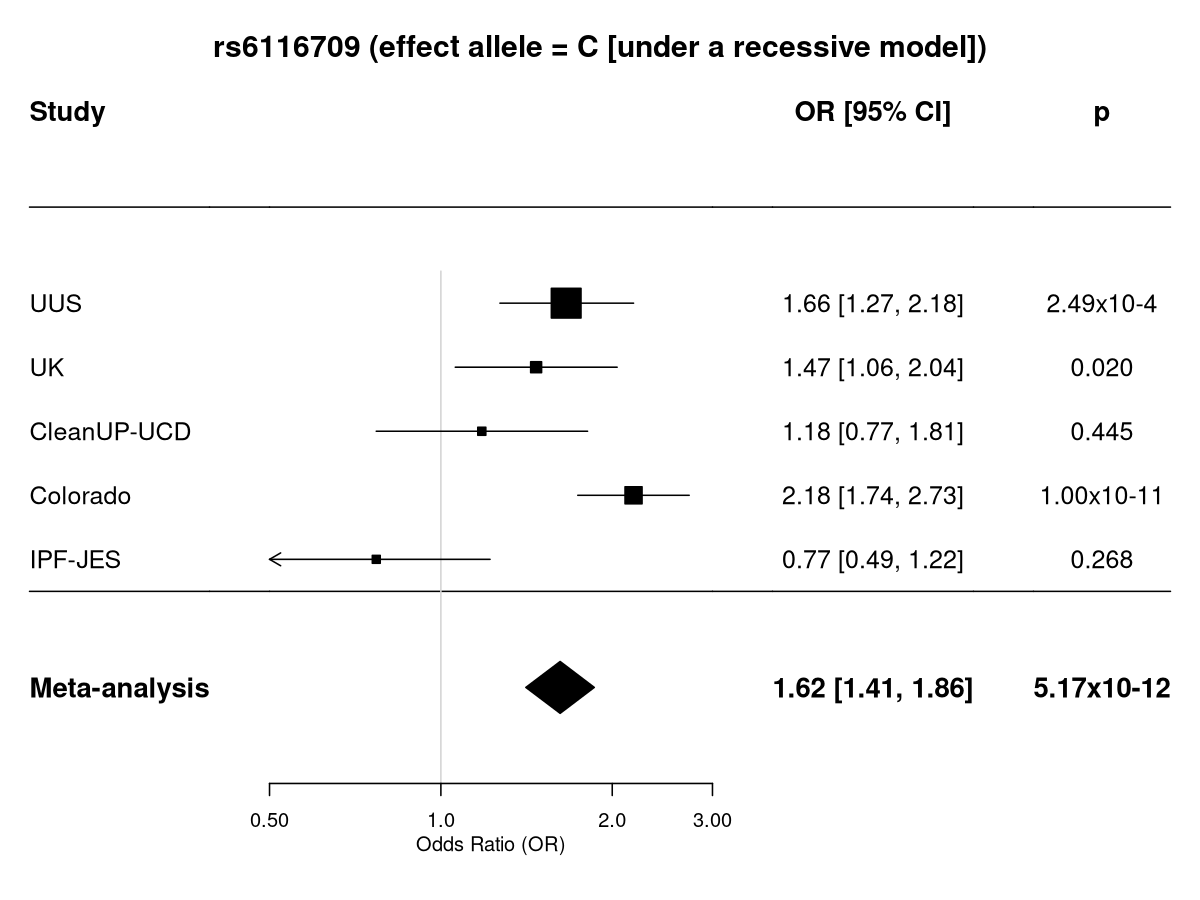

**Figure S3.** Region plots of signals of interest. The y-axis shows the transformed p-values (-log10), while the x-axis represents chromosomal positions (GRCh38). The genome-wide significance threshold (*p*=5x10^-8^) is indicated by the horizontal dashed line. Linkage disequilibrium values (r2) are presented according to the linkage disequilibrium colour scheme of the upper left legend. Plots were generated using LocusZoom (http://locuszoom.org/):

a) Lead variant rs1052053 (*PMF1*)
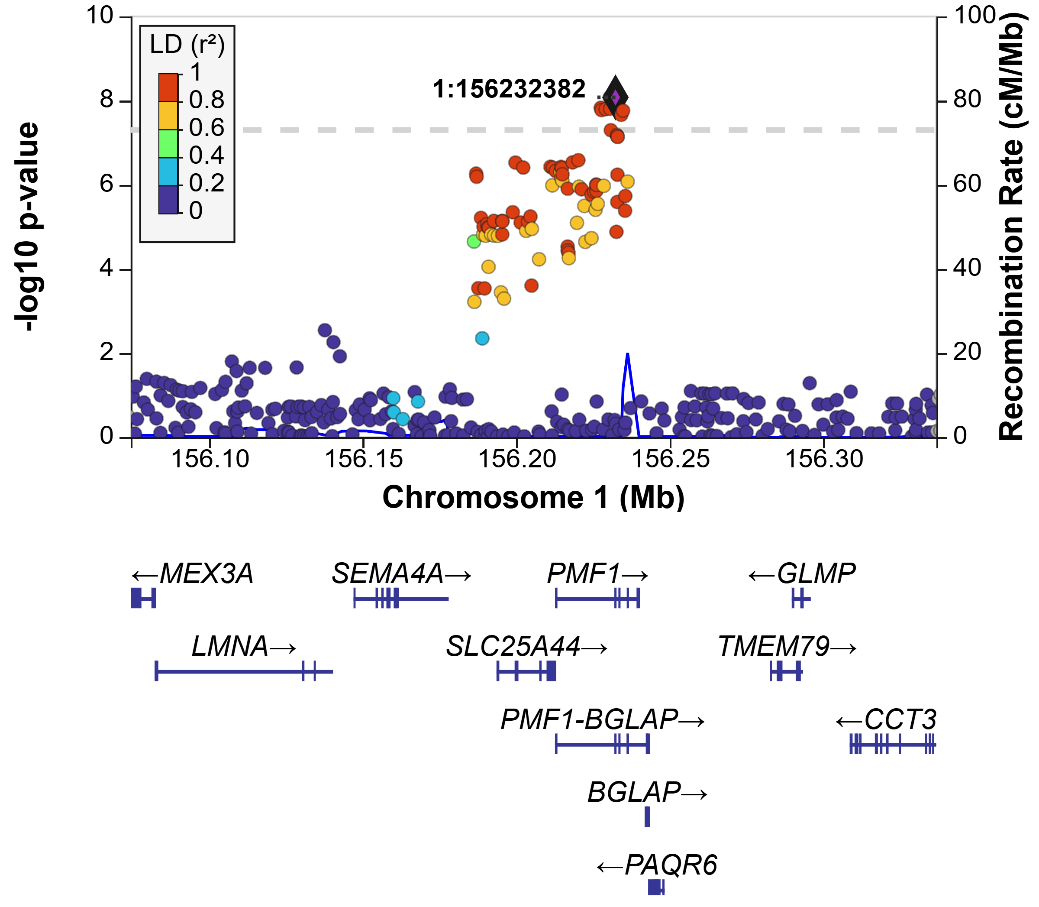

b) Lead variant rs74455724 (*CNTNAP3*)

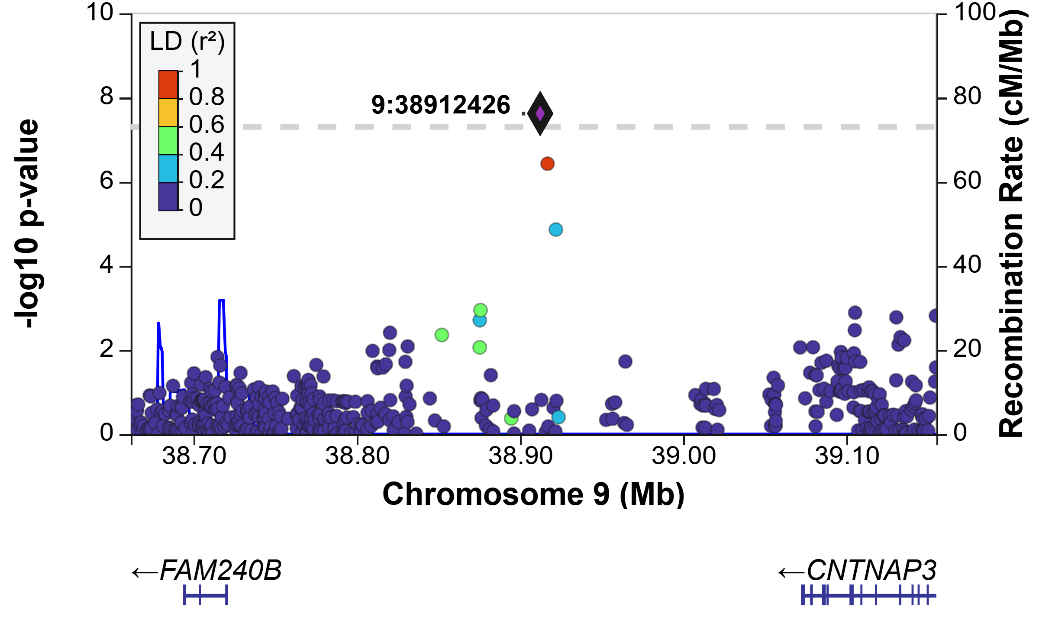

c) Lead variant rs11838956 (*ARHGEF7)*
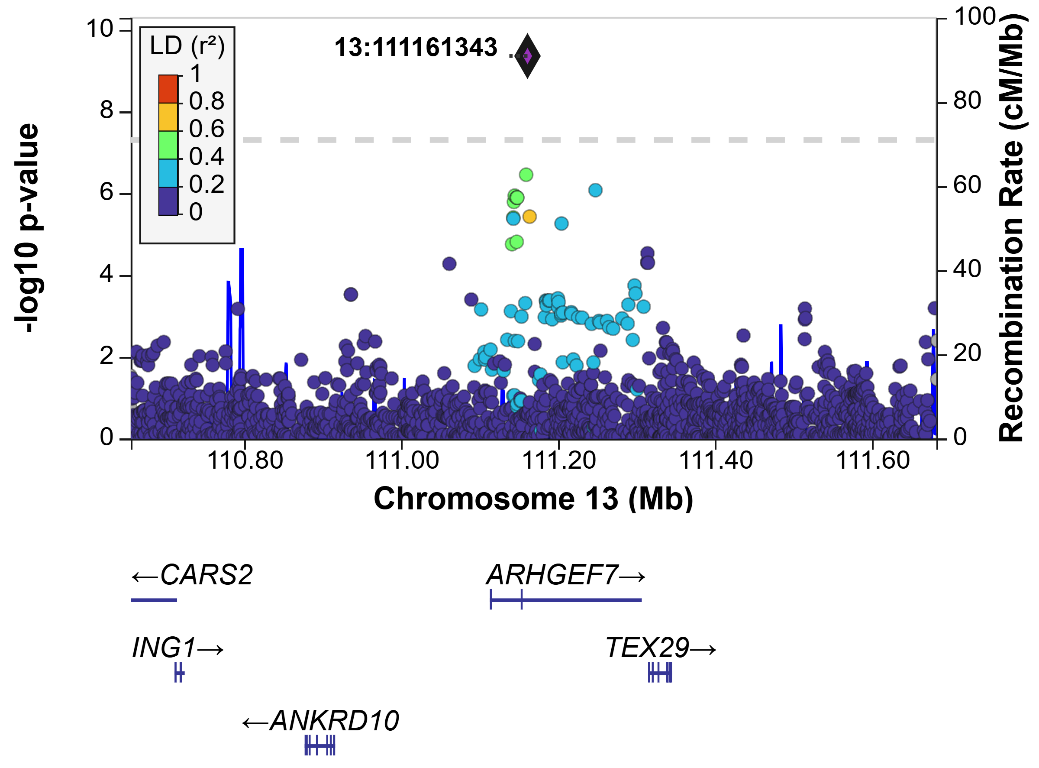

d) Lead variant rs4794159 (*EPN3*)
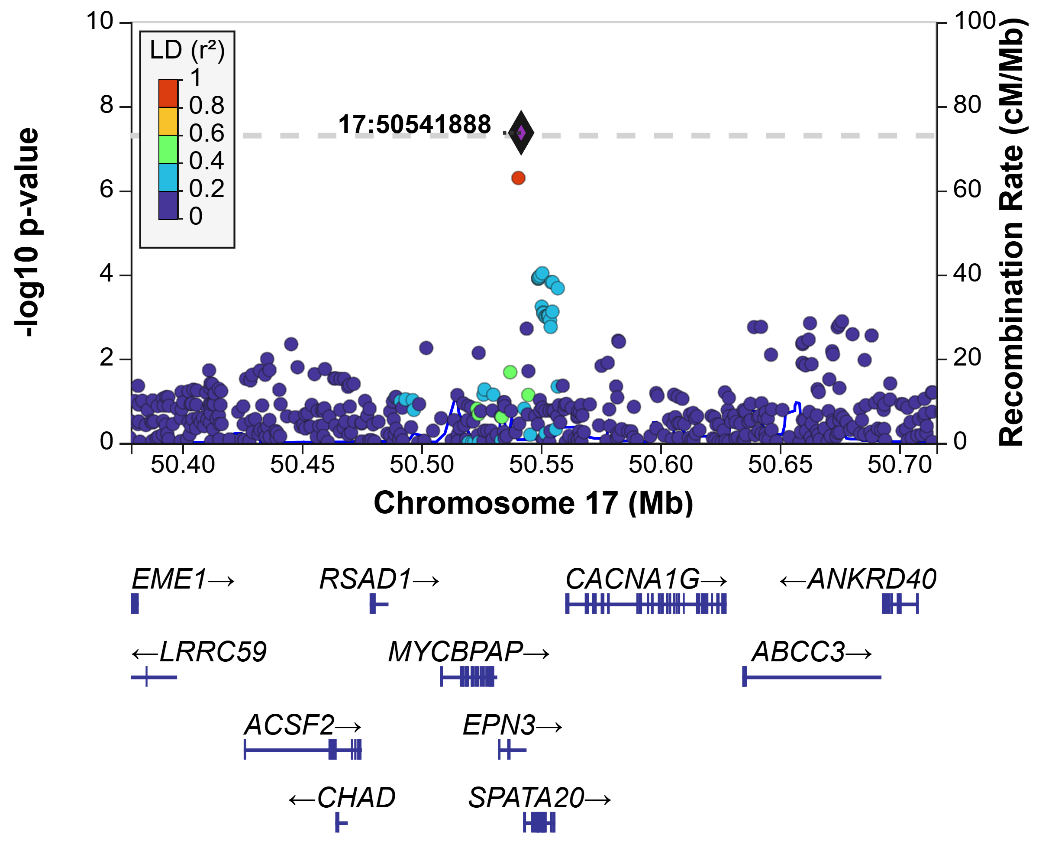

e) Lead variant rs6116709 (*CDS2*)

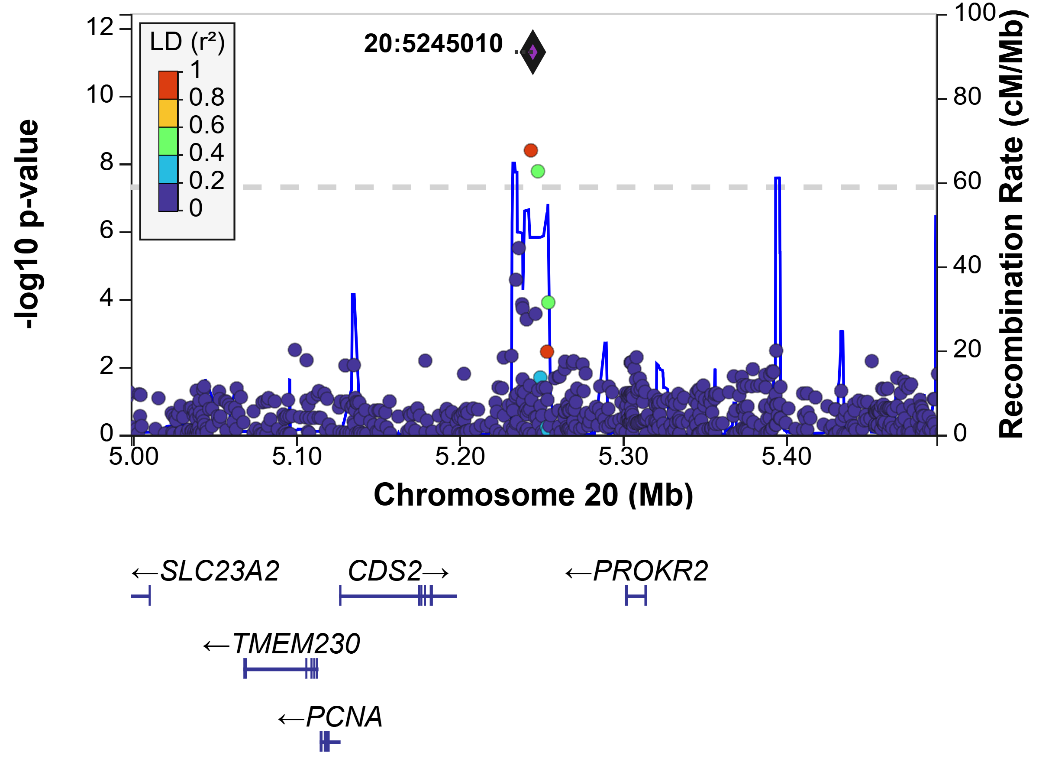

**Figure S4.** Transcripts and tissue expression (from GTEx) of signals that overlap with alternative transcripts (FLIbase_ENIC and/or Ensembl_EIC, which are not categorised as nonsense-mediated mRNA decay [NMD]). The left-hand panel shows gene transcripts with exons (filled according to the exons) and introns (lines). Dark red dashed vertical lines indicate the positions of the SNPs of interest. The right-hand panel is a heatmap showing the average expression of transcripts across tissues. Positive values are shown in a yellow–red–black gradient, while 0 or missing values are shown in white:

a) Highlighting variant in the credible set (chromosome 1, rs2072499, PIP=0.006) that lies within an exon of *SLC25A44* in four transcripts (TCONS_00035947; TCONS_00035943; TCONS_00035940; TCONS_00035935, highlighted in green), with the reference transcript ENST00000359511 based on VEP (TCONS_00035918, highlighted in blue).

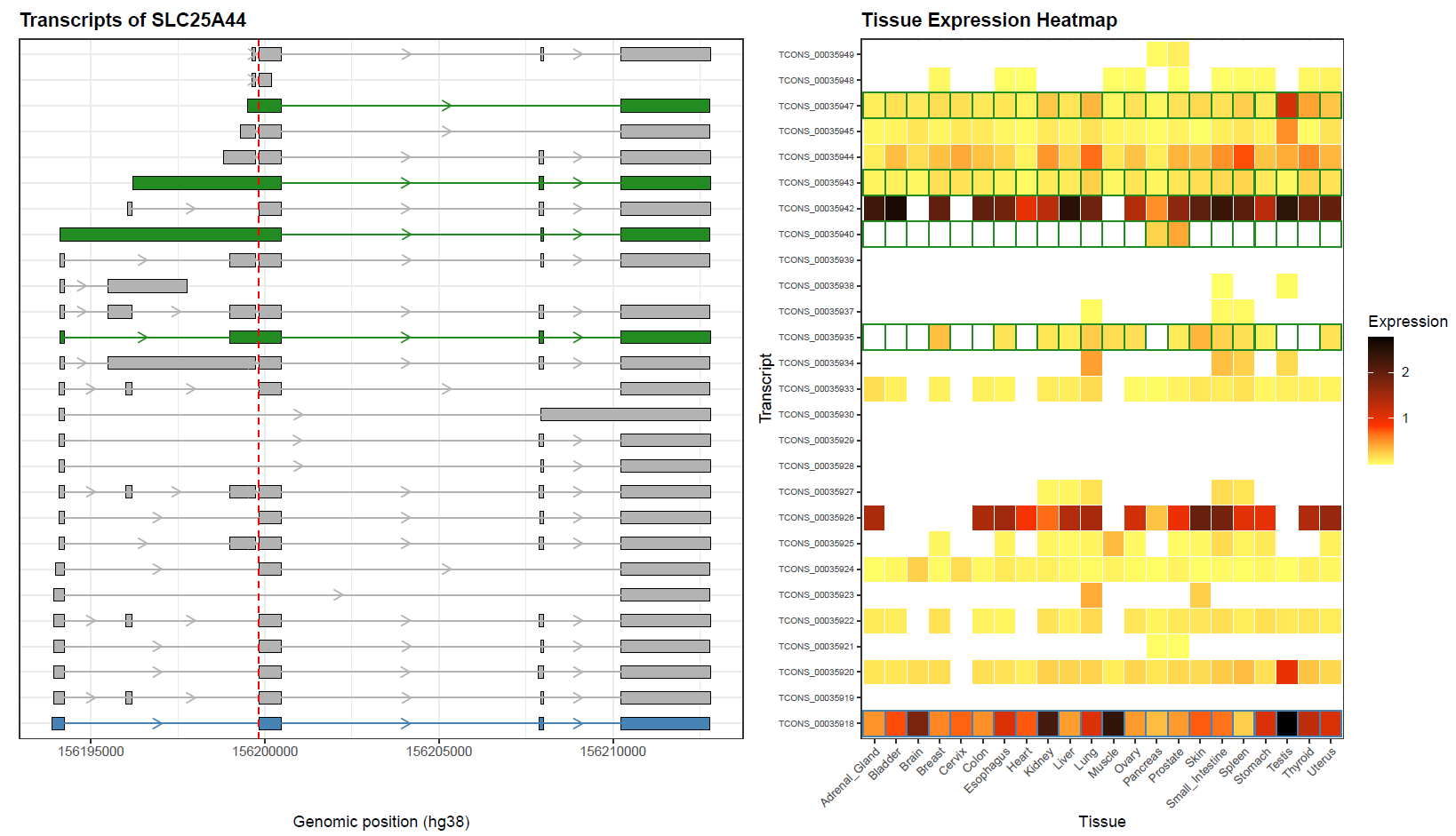

b) Highlighting variant in the credible set (chromosome 17, rs7215760, PIP=0.088) that lies within an exon of *EPN3* in one transcript (TCONS_00434517, highlighted in green), with the reference transcript ENST00000268933.8 based on VEP (TCONS_00434440, highlighted in blue).

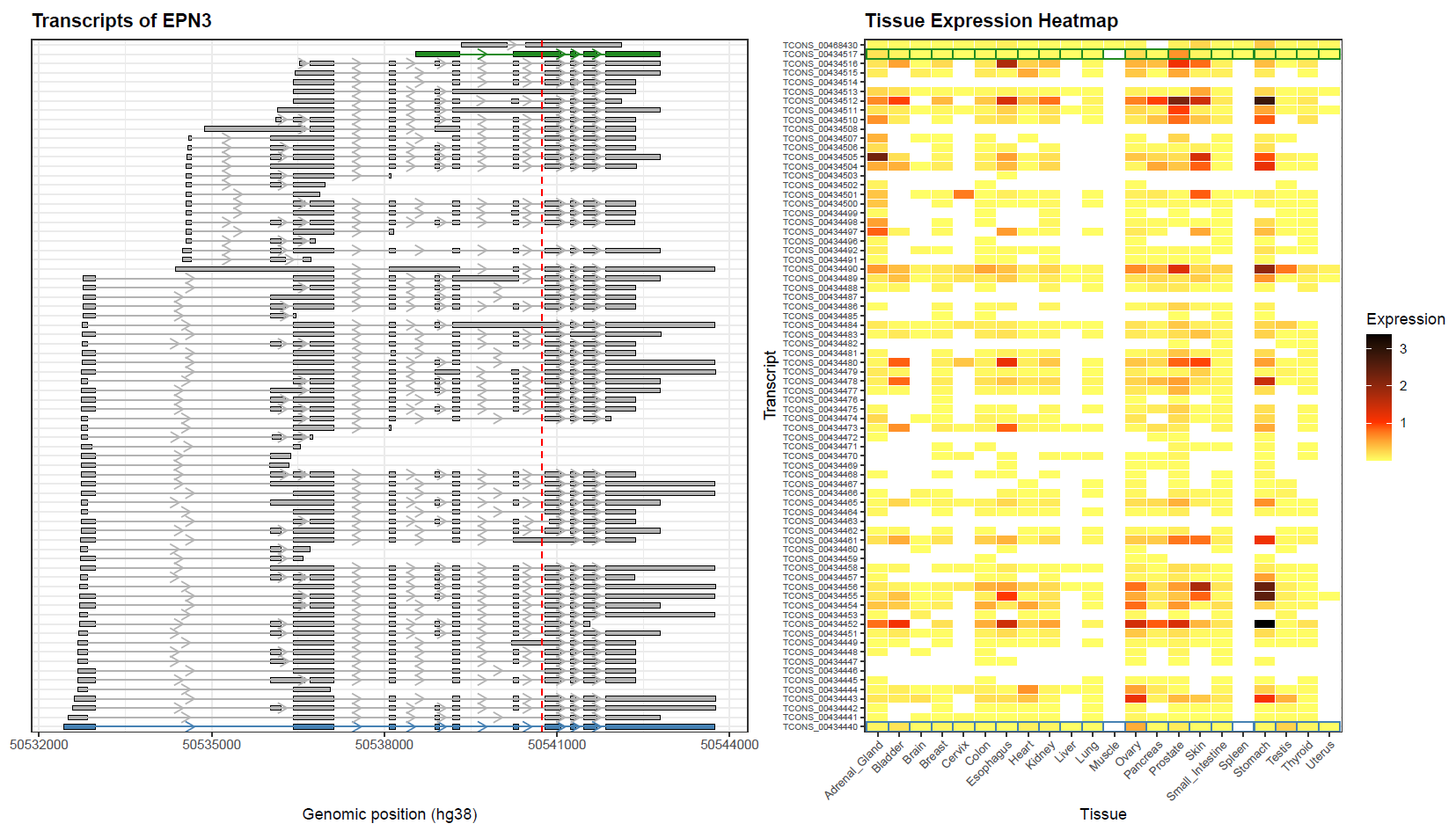

**Figure S5.** Single variant recessive PheWAS results. The PheWAS were aligned to the IPF recessive allele.

a) Lead variant rs1052053-G (*PMF1*)

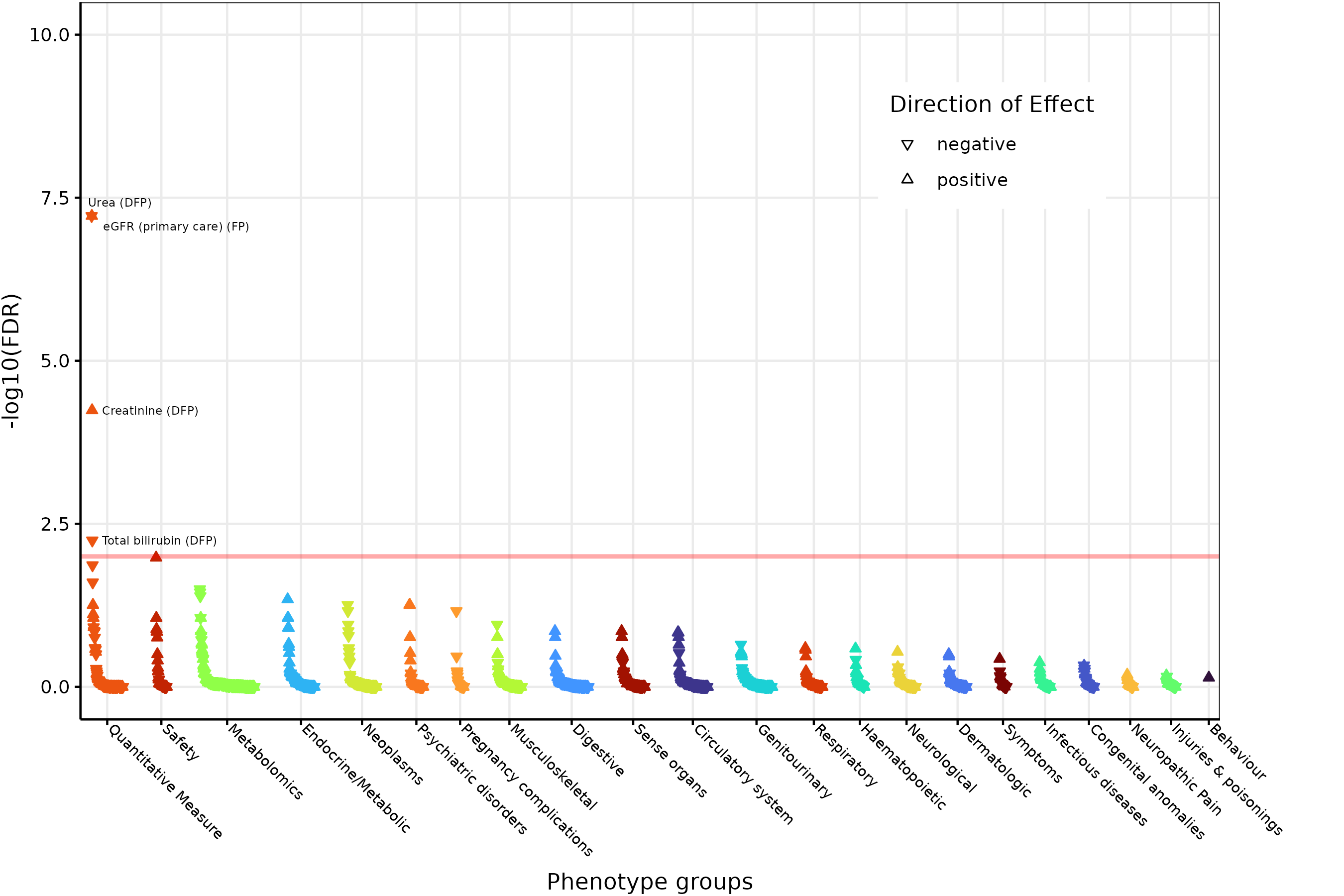

b) Lead variant rs74455724-A (*CNTNAP3*)

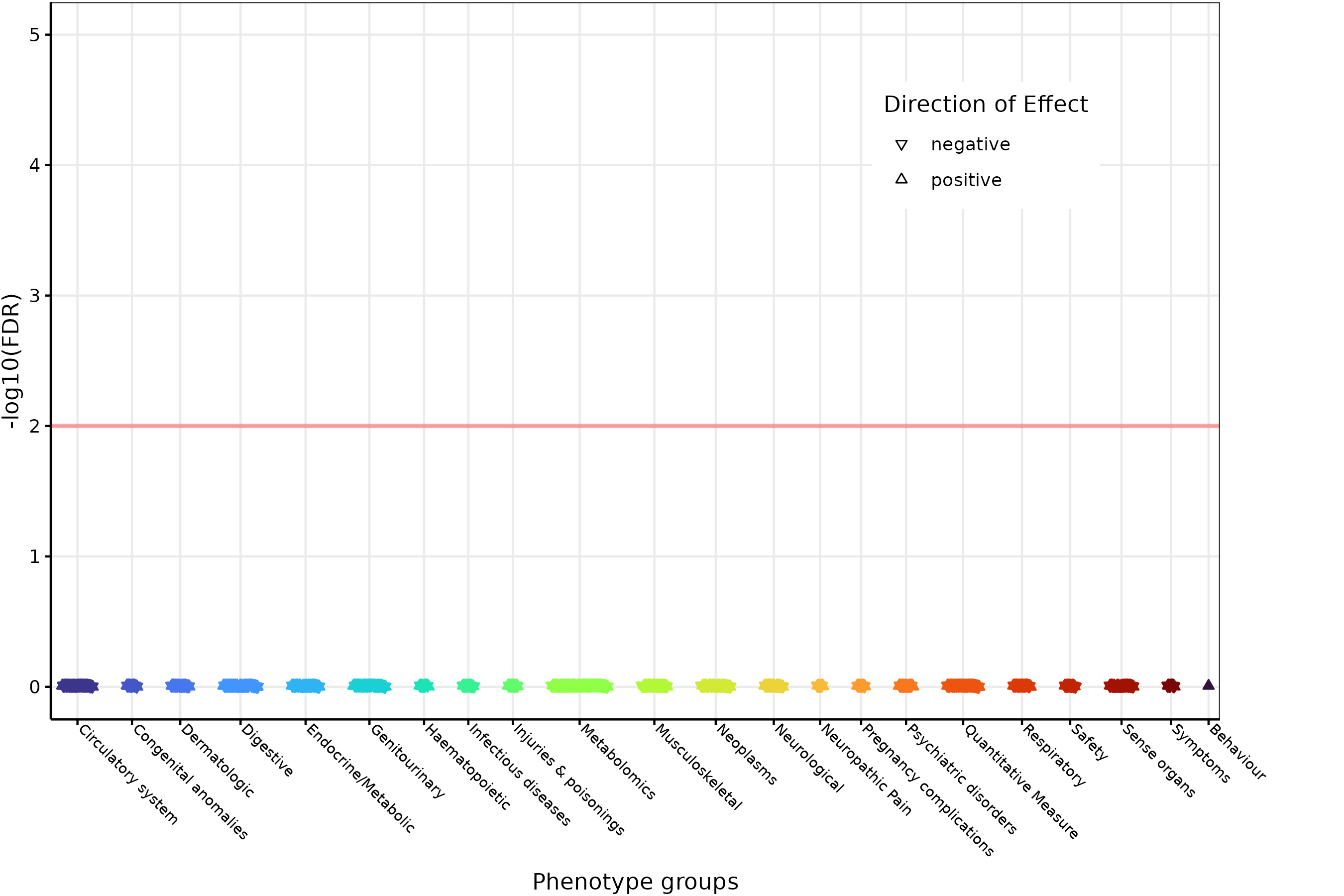

c) Lead variant rs11838956-T (*ARHGEF7)*

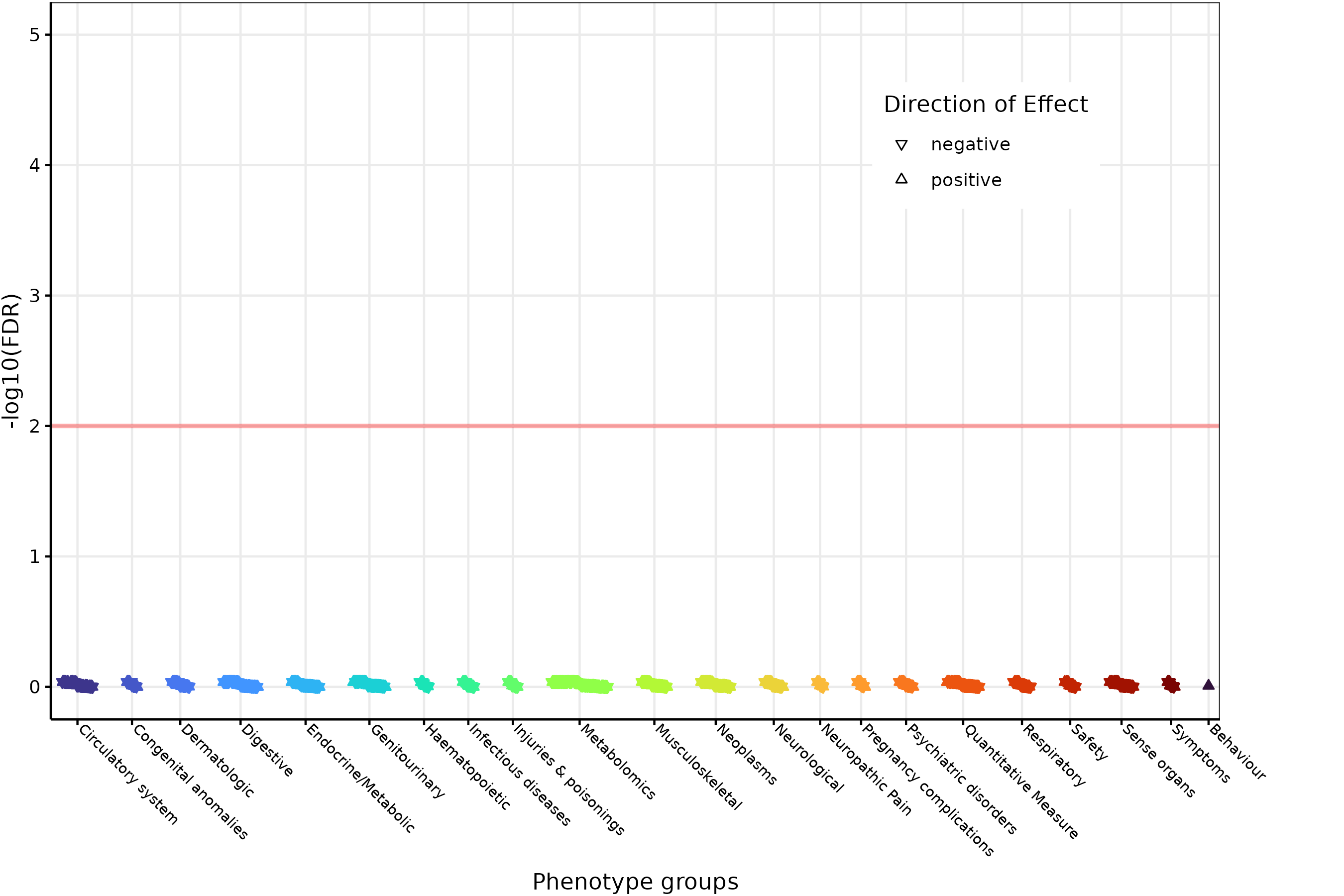

d) Lead variant rs7215760-A (*EPN3*)

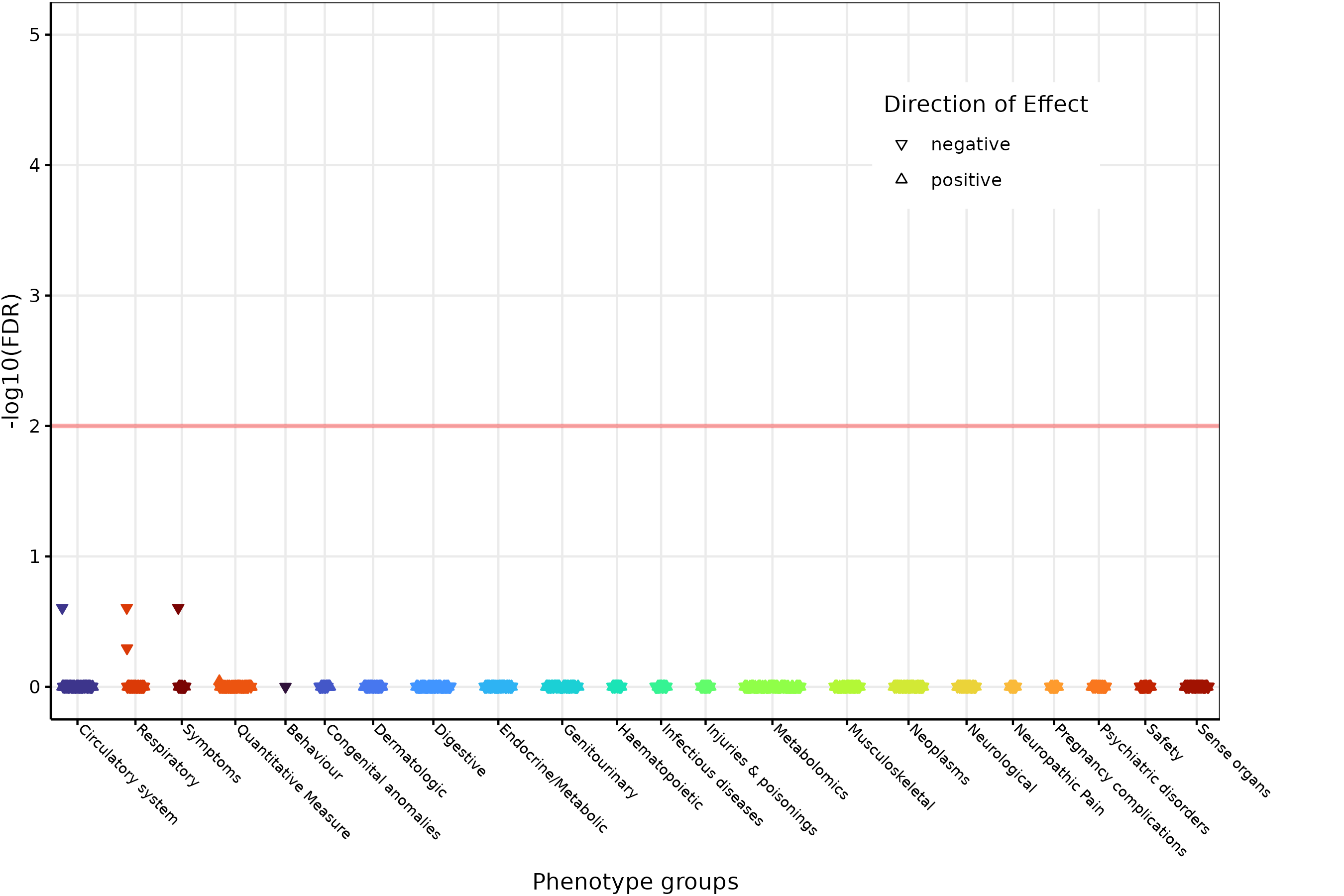

e) Lead variant rs6116709-C (*CDS2*)

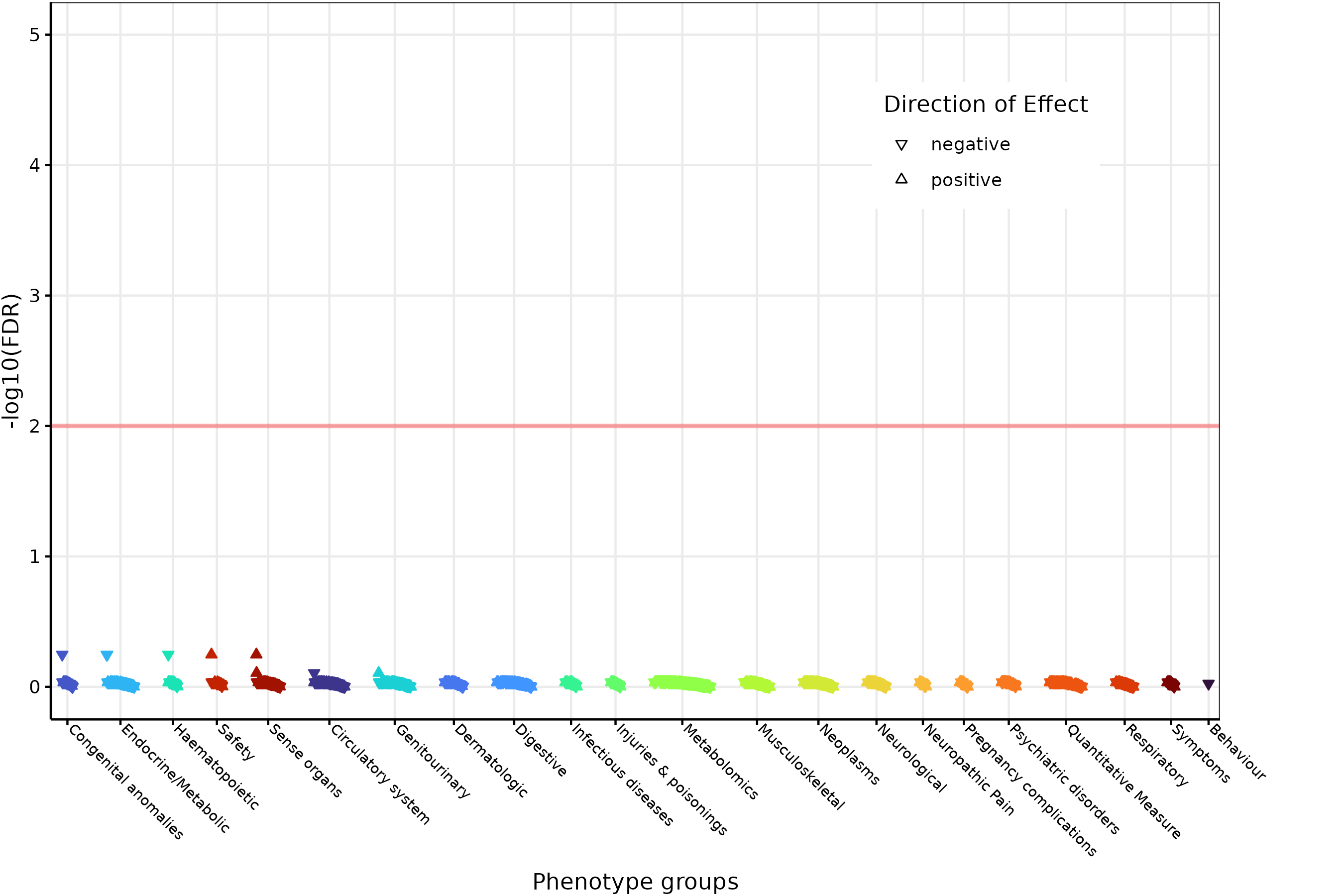

**Figure S6.** Average gene expression in integrated scRNA dataset annotated with HLCA scRNA of: a) PMF1 at HLCA annotation level 3; b) Statistically significant PMF1 expression in AT2, Basal and Fibroblasts; c) ARHGEF7 at HLCA annotation level 3; d) Statistically significant ARHGEF7 expression in AT1, Basal and Fibroblasts; e) EPN3 at HLCA annotation level 3; and f) EPN3 expression in AT1, AT2 and Basal cells (not significant). Statistical significance was calculated using the ‘t.test’ function in R (ggsignif package extension) with a significance threshold of p<0.05.

a) PMF1 at HLCA annotation level 3

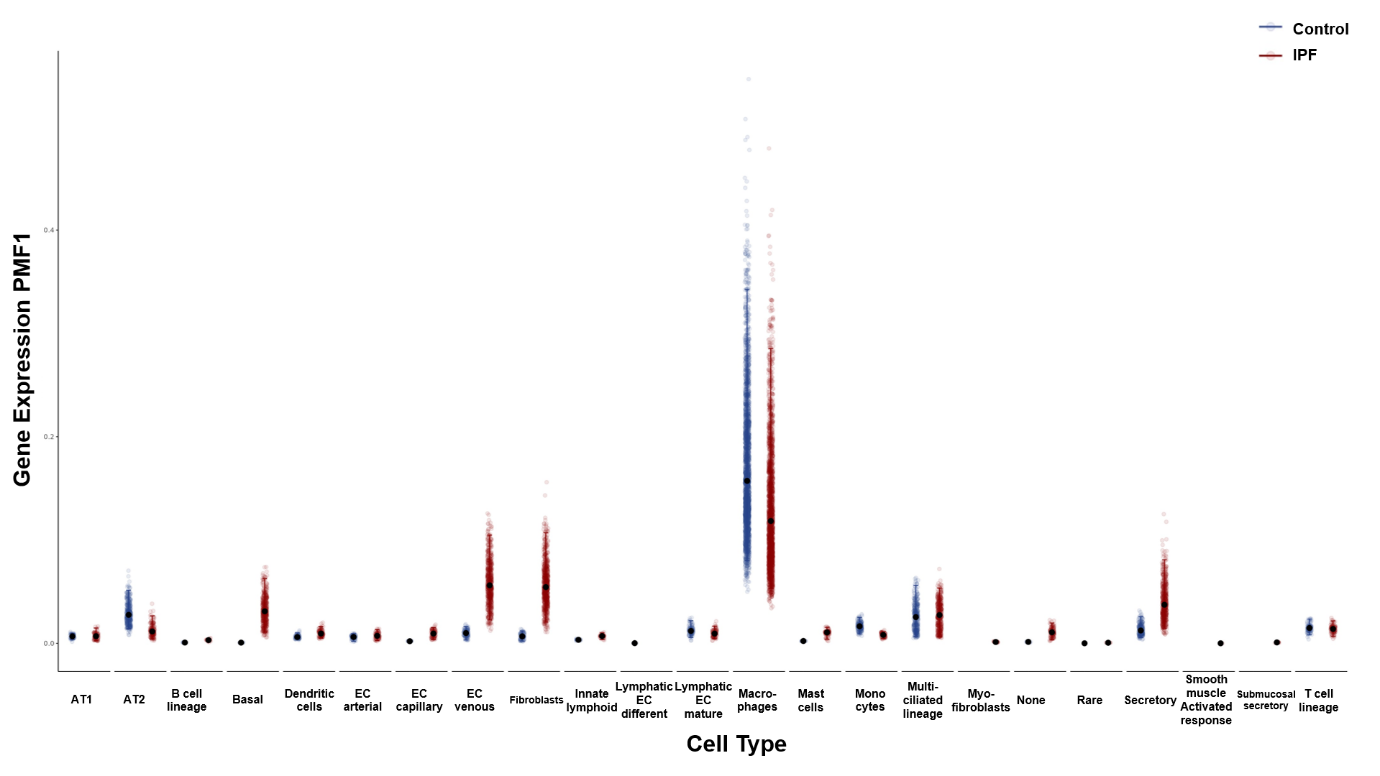

b) Statistically significant PMF1 expression in AT2, Basal and Fibroblasts

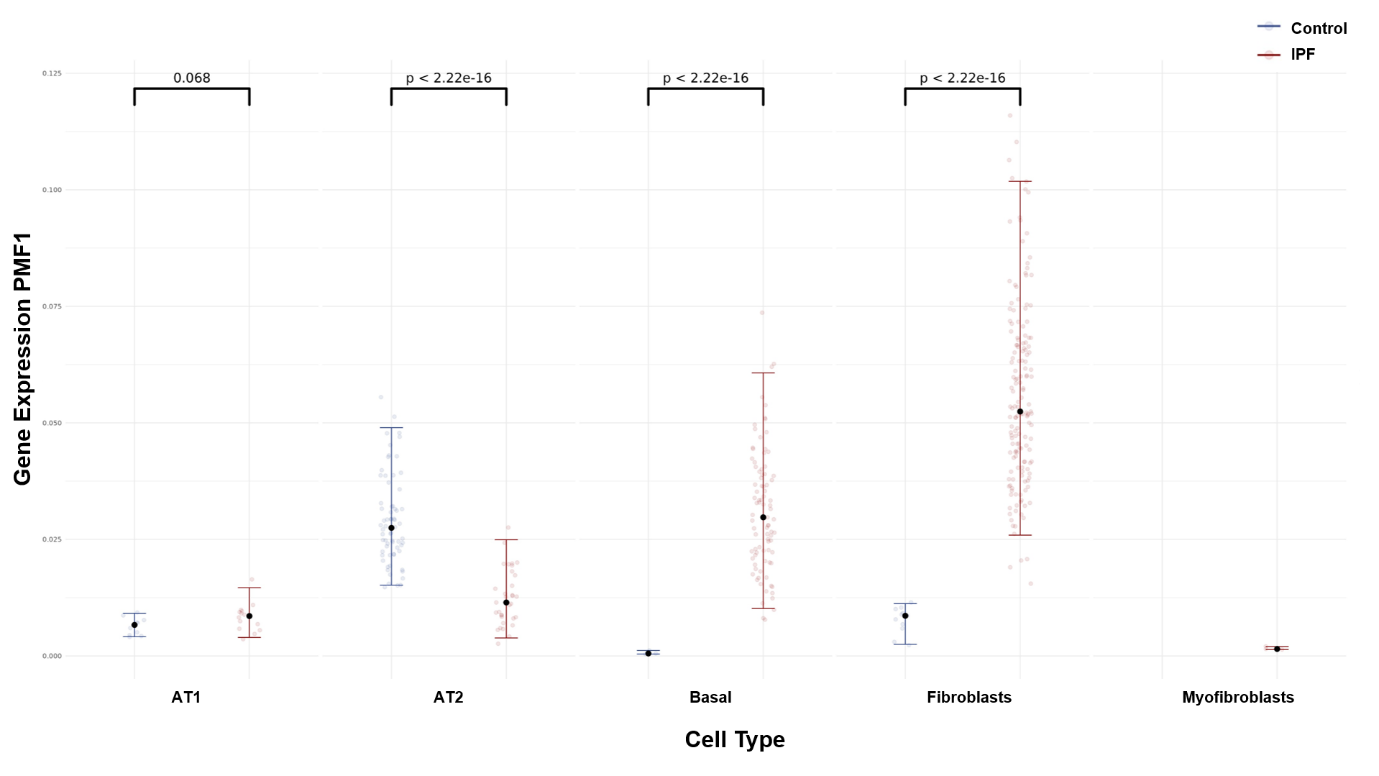

c) ARHGEF7 at HLCA annotation level 3

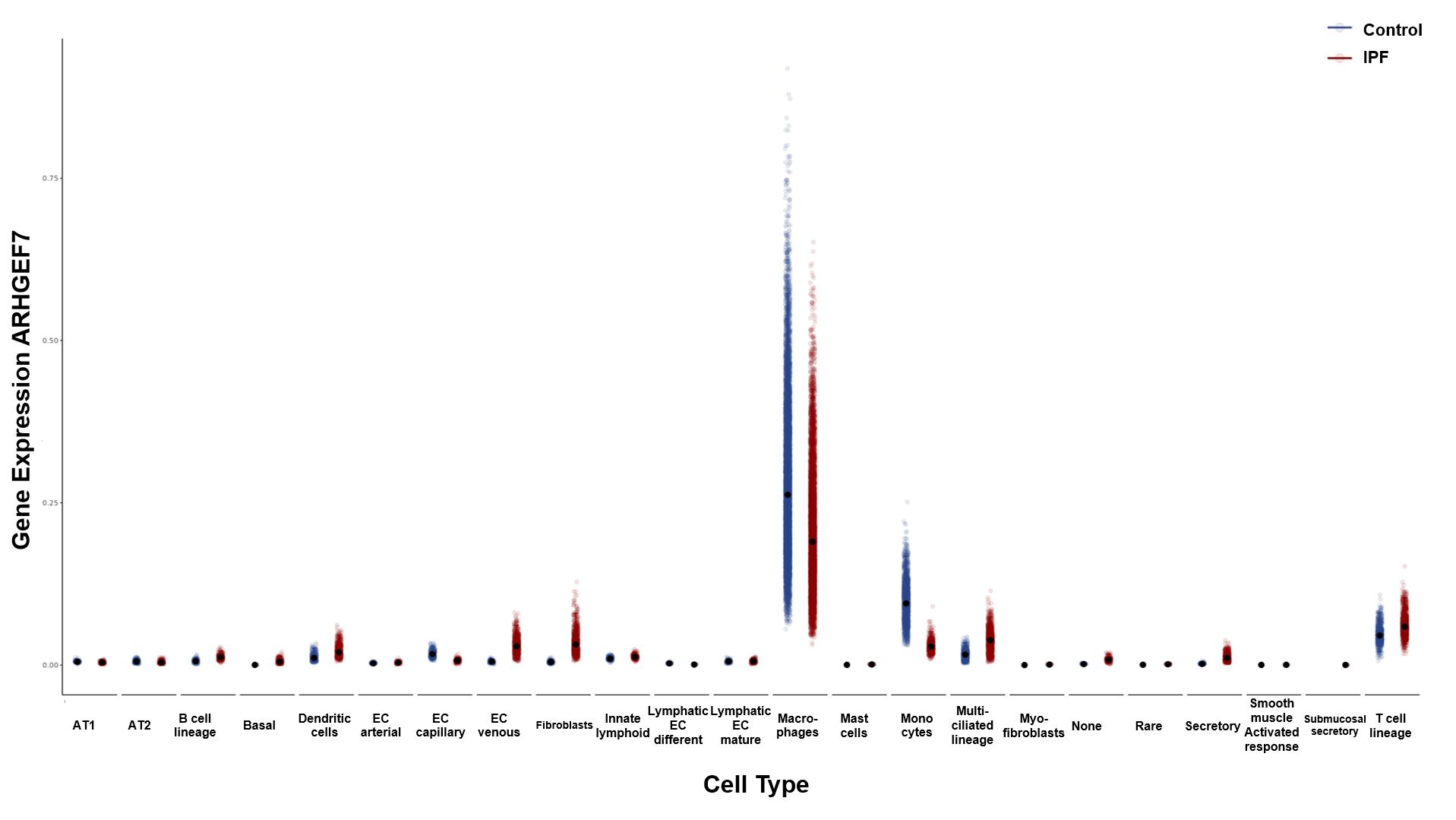

d) Statistically significant ARHGEF7 expression in AT1, Basal and Fibroblasts

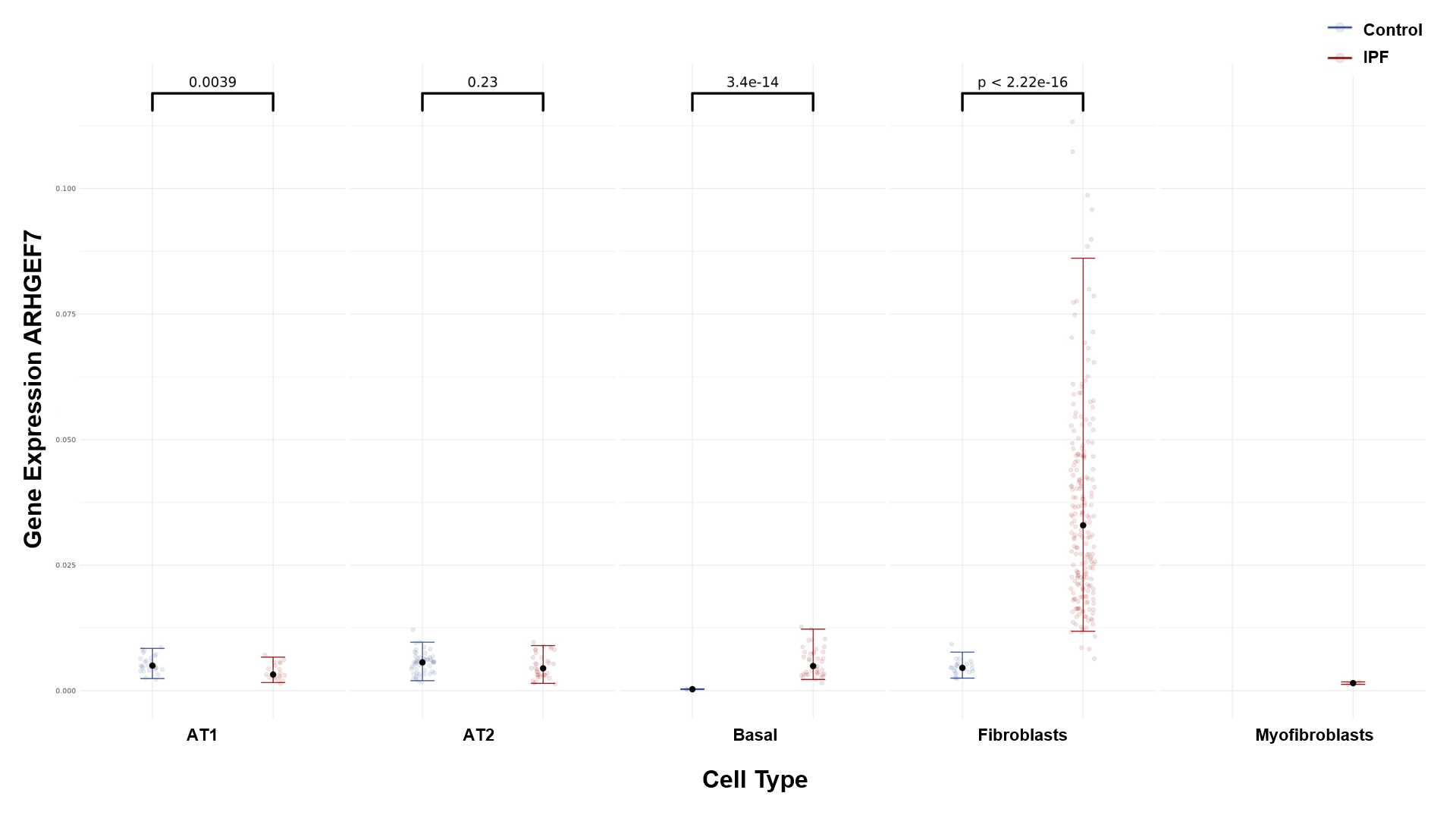

e) EPN3 at HLCA annotation level 3

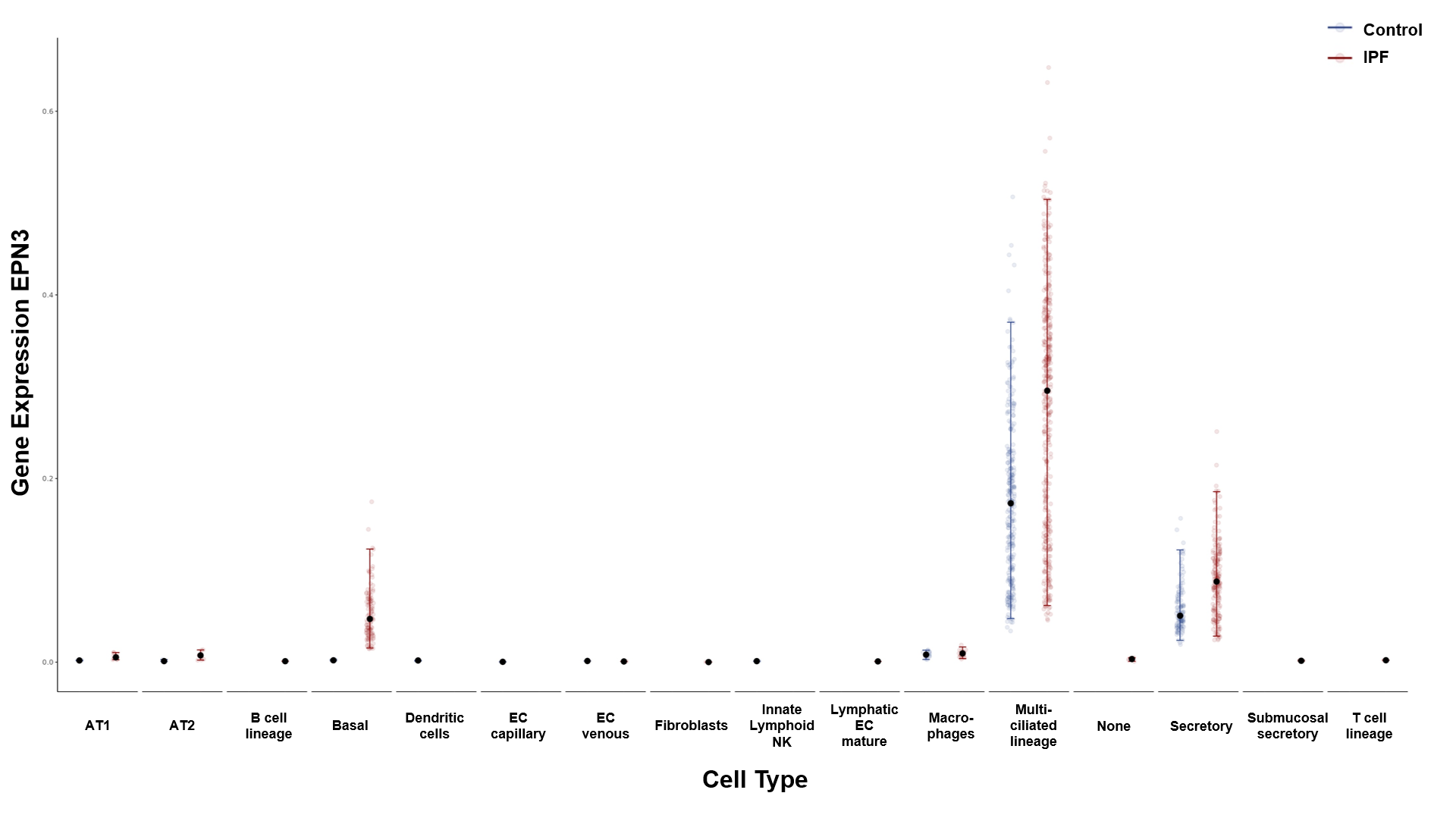

f) EPN3 expression in AT1, AT2 and Basal cells (not significant)

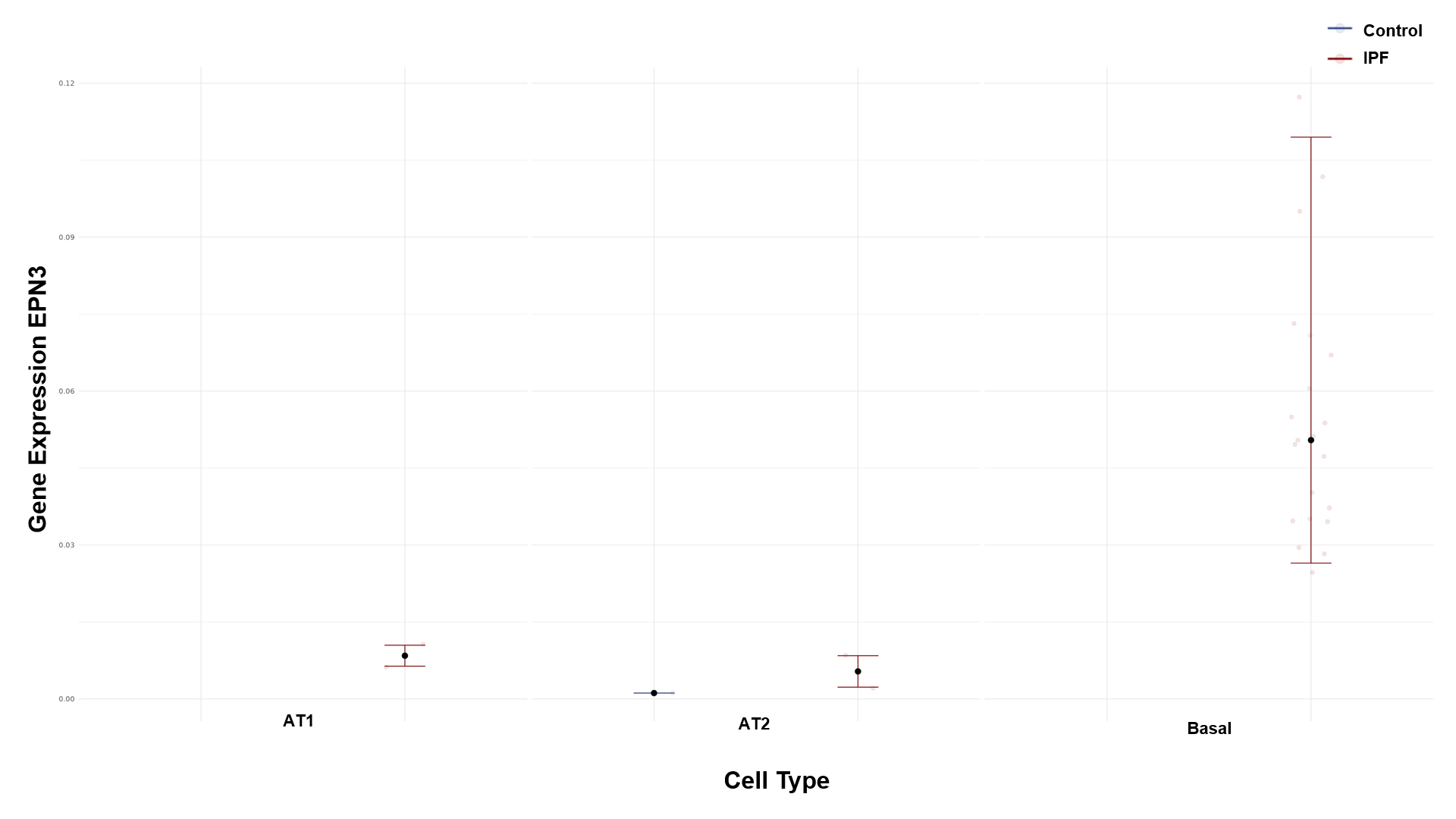

**Figure S7.** mRNA expression of *PMF1* in non-IPF and IPF basal cells (n=3, biological replicates of independent donors). Relative gene expressions of *PMF1* normalised to B2M per sample.

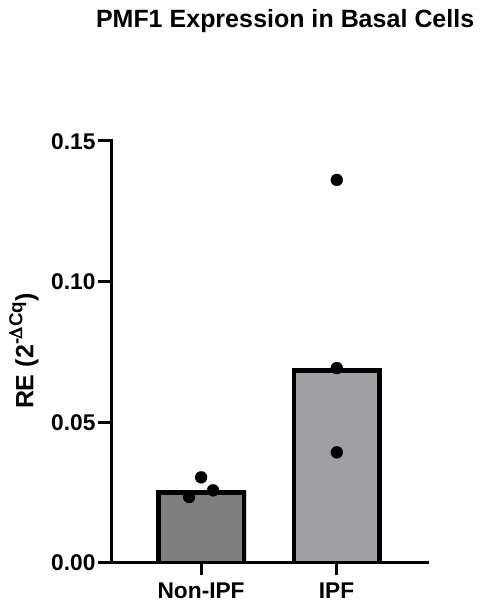

**Figure S8.** Summary plot of the previously reported signals in the non-additive genetic models (dominant and recessive). The x-axis shows the nearby genes harbouring the obtained SNPs. The y-axis shows the transformed p-values (-log10). The horizontal dashed black line corresponds to the genome-wide threshold (*p*=5×10^-8^). Each colour represents the transformed p-value obtained from the different genetic models: in blue the additive results, in red the results of the dominant genetic model and in green the recessive results. rs35705950 (*MUC5B*) SNP is not shown in the plot as it has a *p*<5×10^-120^ in all genetic models.

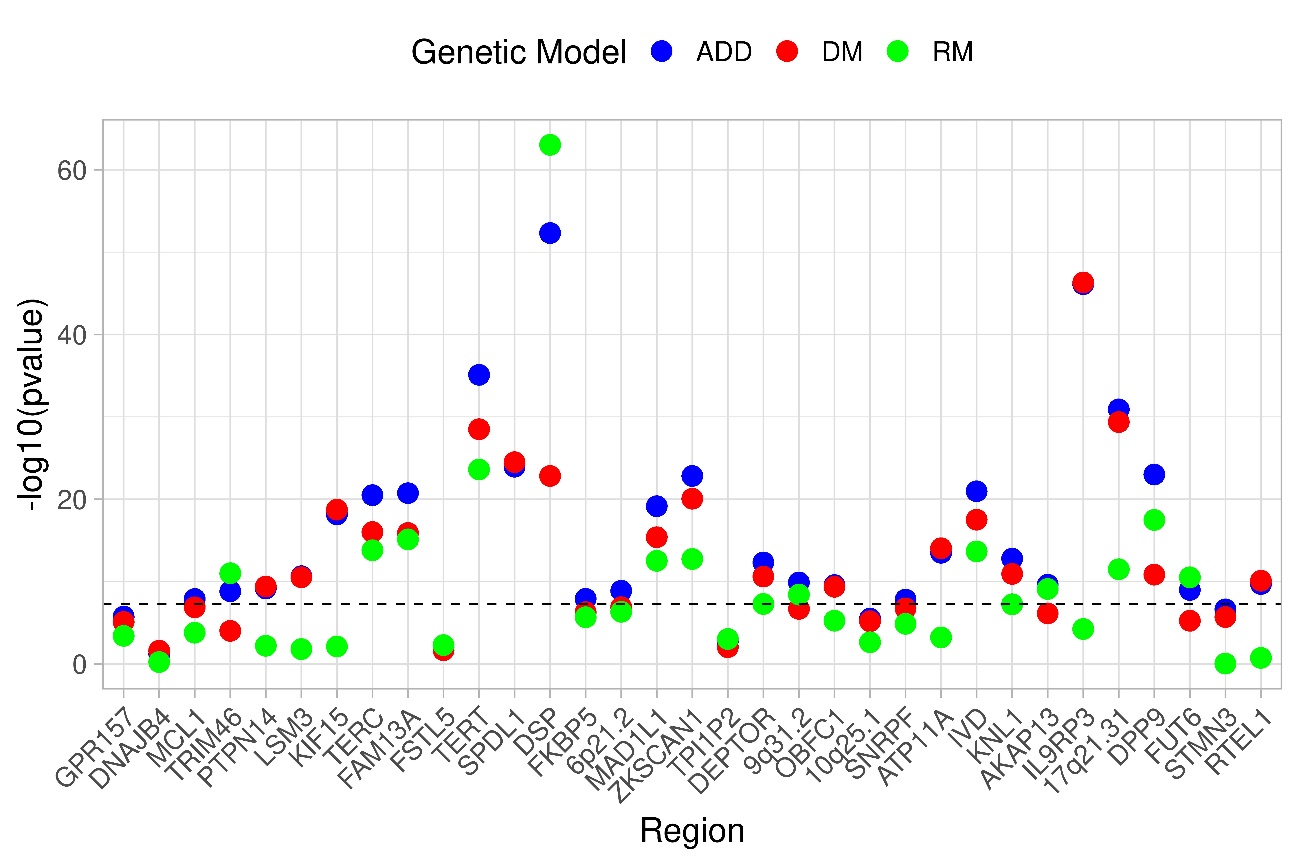
